# A unified framework for cell-type-specific eQTLs prioritization by integrating bulk and scRNA-seq data

**DOI:** 10.1101/2024.05.27.24307972

**Authors:** Xinyi Yu, Xianghong Hu, Xiaomeng Wan, Zhiyong Zhang, Xiang Wan, Mingxuan Cai, Tianwei Yu, Jiashun Xiao

## Abstract

Genome-wide association studies (GWASs) have identified numerous genetic variants associated with complex traits, yet the biological interpretation remains challenging, especially for variants in non-coding regions. Expression quantitative trait loci (eQTLs) studies have linked these variations to gene expression, aiding in identifying genes involved in disease mechanisms. Traditional eQTL analyses using bulk RNA sequencing (bulk RNA-seq) provide tissue-level insights but suffer from signal loss and distortion due to unaddressed cellular heterogeneity. Recently, single-cell RNA sequencing (scRNA-seq) has provided higher resolution enabling cell-type-specific eQTL (ct-eQTL) analyses. However, these studies are limited by their smaller sample sizes and technical constraints. In this paper, we present a novel statistical framework, IBSEP, which integrates bulk RNA-seq and scRNA-seq data for enhanced ct-eQTLs prioritization. Our method employs a Bayesian hierarchical model to combine summary statistics from both data types, overcoming the limitations while leveraging the advantages associated with each technique. Through extensive simulations and real-data analyses, including peripheral blood mononuclear cells and brain cortex datasets, IBSEP demonstrated superior performance in identifying ct-eQTLs compared to existing methods. Our approach unveils new transcriptional regulatory mechanisms specific to cell types, offering deeper insights into the genetic basis of complex diseases at a cellular resolution.

## Introduction

Genome-wide association studies (GWASs) have made significant achievements in identifying genomic regions associated with complex traits/diseases, yet the biological interpretation of GWAS results remains challenging as the majority of risk variants located in non-coding regions of the genome [1]. Among numerous studies annotating genetic variations, expression quantitative trait loci (eQTLs) studies link the risk variations to their potentially regulating target genes, aiding in identifying genes that may participate in disease mechanisms [2]. Studies have shown that genetic variations associated with complex traits are enriched in identified eQTLs [3]. Given the significant relevance of eQTLs studies in interpreting GWAS results, researchers worldwide have accumulated substantial gene expression and genotype data for eQTLs studies [4, 5], laying the foundation for uncovering the genetic basis of complex traits [6].

So far, most eQTLs studies have been based on bulk RNA sequencing (bulk RNA-seq) at the tissue level, measuring the average gene expression levels from hundreds to millions of cells within a tissue. One of the most prominent eQTLs studies comes from the Genotype-Tissue Expression (GTEx) project, which analyzed 15,201 RNA sequencing samples from 49 tissues of 838 donors, identifying tissue level eQTLs in 94.7% of protein-coding genes [4]. Despite the fruitful discoveris of tissue level eQTLs, they still only explain a moderate fraction of GWAS signals [7]. In recent years, increasing evidence suggests that transcriptional regulation mechanisms are dynamic and highly dependent on cell type or cell state [8]. Some eQTLs have been demonstrated to be detectable only in specific cell types[8], indicating the critical importance of cell-type-specific eQTLs (ct-eQTLs) prioritization for better interpreting and understanding GWAS results.

Studies of ct-eQTLs can be divided into two categories: those based on traditional bulk RNA-seq data and those based on single-cell RNA sequencing (scRNA-seq) data. Given the abundant bulk RNA-seq data, ct-eQTLs can be identified by detecting the interaction effects of single-nucleotide polymorphisms (SNPs) and cell type proportions on gene expression, known as interaction QTLs (ieQTLs). For instance, Zhernakova et al [9] introduces a “genotype-cell type proportion” interaction term into the eQTL linear model and observed that 12% of cis-regulated genes showed context-dependent eQTL effects. Further studies include Decon-eQTL [10] that included multiple cell types in the interaction term, and CSeQTL [11] that models gene expression using binomial or Poisson distribution. While this approach can be directly applied to existing bulk RNA-seq data, its statistical power is unsatisfactory, particularly when locating eQTLs specific to rare cell types. The other type of approach directly utilizes scRNA-seq data to locate ct-eQTLs. For instance, Van et al analyzed peripheral blood mononuclear cells (PBMCs) scRNA-seq data from 45 donors and identified a total of 379 eQTLs across six cell types, including 48 newly discovered ct-eQTLs [12]; Bryois et al used brain cortex scRNA-seq data from 192 individuals and identified a total of 7,607 eQTLs across eight major brain cell types, with 46% showing cell-type-specific associations, particularly in microglia cells [13]. However, due to the high cost and technical constraints (e.g dropouts) of scRNA-seq, these studies typically have small-scale and low-quality gene expression counts, limiting the power of ct-eQTLs mapping. In summary, tissue-level sequencing data has a larger sample size but low resolution, while single-cell-level sequencing data has high resolution but a smaller scale.

Although the above methods can be utilized for ct-eQTLs mapping, they are not yet perfect in terms of effectiveness, applicability, and scalability. On the one hand, both bulk RNA-seq and scRNA-seq based ct-eQTLs studies are currently conducted independently, without finding effective methods to overcome the heterogeneity between the two types of data and integrate them into a unified model framework [14]. On the other hand, existing ct-eQTLs methods requires individual-level genotype and transcriptomic data, thus cannot fully make use of widely available summary statistics[9, 10, 11, 15, 16]. Besides, current ct-eQTLs analysis ignored the widespread genetic correlations among cell types, especially biologically close ones, and performed ct-eQTLs mapping on a single cell type at a time, leading to a suboptimal statistical power [13, 17].

In this paper, we develop a unified statistical framework for integrating bulk RNA-seq and scRNA-seq data for ct-eQTLs prioritization (IBSEP). We first demonstrate the heterogeneity of bulk RNA-seq and scRNA-seq data can be overcome with theoretical justification, and then use a Bayesian hierarchical linear model to link these two types of data. The keys to the success of IBSEP are three-fold. First, by integrating tissue-level and cell-type-level eQTL data into a unified model, it can effectively leverage the advantages of each type of data to enhance the statistical power of ct-eQTLs prioritization. Second, by directly modeling the eQTLs summary statistics, it can use these summary statistics as input, enabling greater utility in large-scale data integration analyses. Third, by incorporating the cis-coheritability between SNPs across cell types, it can consider the complex structural relationships between cell types, revealing both shared and specific patterns of transcriptional regulation across different cell types. Through comprehensive simulation studies, we demonstrated that IBSEP outputs well-calibrated *p*-values and achieves significant power gains. Then, by separately integrating a PBMCs [18] and a brain [13] scRNA-seq dataset with corresponding GTEx bulk RNA-seq datasets [4], IBSEP unveiled more ct-eQTLs and the underlying cell type-specific transcriptional regulatory mechanisms. Further colocalization analysis of GWAS risk SNPs and IBSEP ct-eQTLs discovered novel target genes mediating immune and brain disease associations. These findings provide new insights for uncovering the underlying mechanisms of gene transcription regulation associated with diseases at the resolution of cell types.

## Results

### Method overview

Current independent ct-eQTLs studies overlook the inherent correlation between bulk RNA-seq and scRNA-seq data, leading to large room for improvement in statistical power for ct-eQTLs prioritization. IBSEP addresses the challenges of ct-eQTLs prioritization from a new perspective (Fig. 1). Essentially, bulk level gene expression of a tissue sample can be viewed as a weighted average of gene expression at the cell type level, with the weights being the cell type proportions of the tissue sample. Given this fact, we designed a unified statistical framework to integrate bulk RNA-seq and scRNA-seq data. Despite cell type proportion variation in tissue samples, we provide theoretical justification and empirical evidence to demonstrate that only the average cell type proportions of tissue samples are required, which enables us to use summary statistics instead of individual-level data. To our knowledge, our approach IBSEP is the first method to demonstrate that the integration of bulk RNA-seq and scRNA-seq data is feasible with summary statistics, and the first method that effectively leverages the advantages of each type of data to enhance the statistical power for ct-eQTLs prioritization.

**Figure 1.**
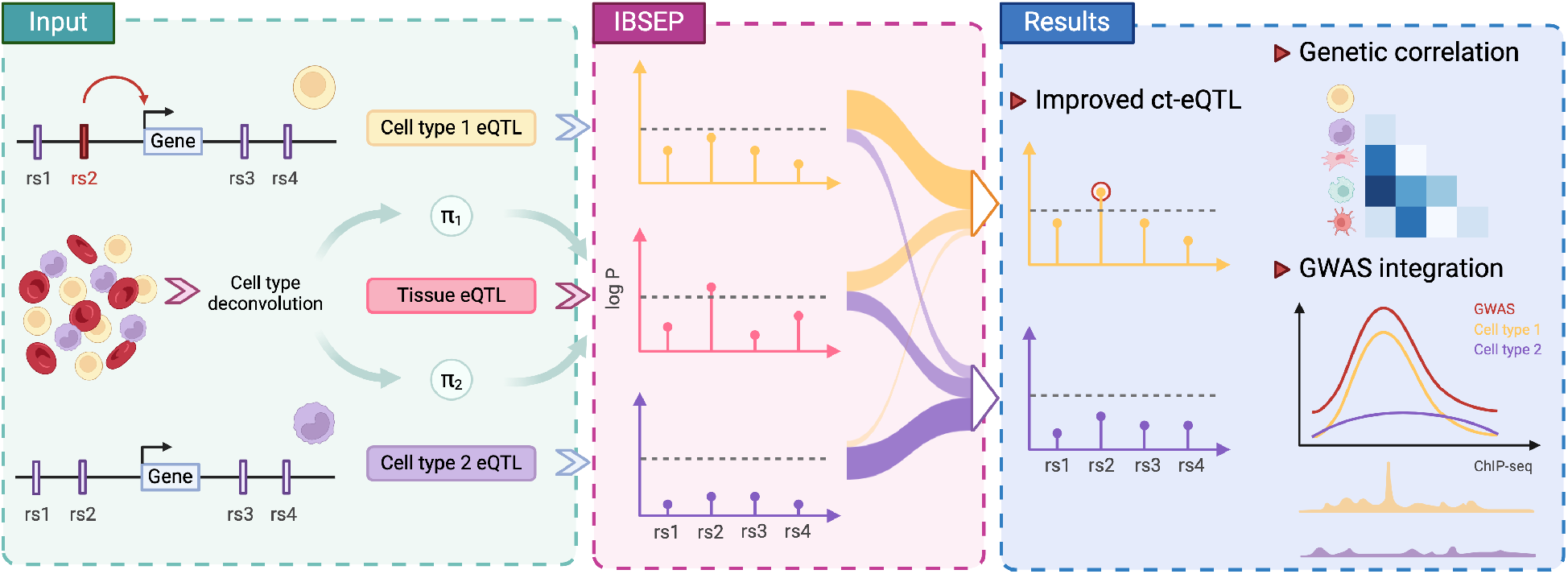
Overview of IBSEP. IBSEP is designed to improve ct-eQTLs prioritization by integrating bulk RNA-seq and scRNA-seq data, revealing the heterogeneity of transcriptional regulation among different cell types. The workflow of IBSEP begins by estimating the cell type proportions for bulk RNA-seq samples. Then, it takes summary statistics of cell type-level eQTLs from scRNA-seq data, tissue-level eQTLs from bulk RNA-seq data and the estimated cell type proportions as input to a Bayesian hierarchical linear model and output improved ct-eQTLs summary statistics. Finally, by integrating with disease GWAS results using colocalization analysis, the improved ct-eQTLs results can provide a comprehensive and in-depth understanding of the genetic basis and pathogenic mechanisms of complex diseases at the resolution of cell type.

In short, IBSEP takes summary statistics of ct-eQTLs from scRNA-seq data with a smaller sample size, summary statistics of tissue-level eQTLs from bulk RNA-seq data with a larger sample size, and the average cell type proportion in tissue samples as model inputs, and output improved ct-eQTLs summary statistics. Details of the model are described in the Methods section. IBSEP significantly enhances the statistical power of ct-eQTLs mappings while producing well-controlled type I errors. With our model design innovations, IBSEP shows high computational efficiency by offering a closed-form solution at each step. Moreover, the output of IBSEP can be readily applied in downstream analyses, such as colocalization with GWAS risk variants, to more effectively discover gene regulation underlying the disease mechanisms at the cell type level.

### Simulation study

One of the key innovations of IBSEP is the theoretical justification that only the average cell type proportions of tissue samples are required in the integration analysis. We conduct simulations to verify this approximation via using real genotypes from UK Biobank (UKBB) individuals. We compared the marginal effect sizes estimates calculated from tissue models with individual 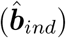and average cell type proportions 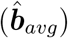 under different samples sizes and cell type proportion variances (More details are provided in the Method section). Consistent with the theoretical result, the correlation between 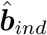 and 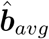 was already high with a small sample size (e.g *N* = 100) and it approached 1 as sample size grows. Besides, 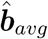 became closer to 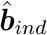 with smaller cell type proportion variance (Fig. S1).

After validating the foundation of our model, we conducted a series of experiments to evaluate the performance of IBSEP. Firstly, we examined whether IBSEP could control type I errors under different scenarios. For simplicity but without loss of generality, we only considered two cell types (cell type 1 and cell type 2). We randomly selected 100 samples from the UKBB as the genotypes for scRNA-seq data (**X**_*c*_) and another 1000 samples for bulk RNA-seq data (**X**_*t*_). These samples were taken from the first linkage disequilibrium (LD) window (containing *M* = 275 SNPs) on chromosome 20. We treated these 275 SNPs as local SNPs mapped to a target gene and performed eQTL analysis. We assumed that there were no cis-SNPs (SNPs with non-zero effects) in cell type 1 and we randomly selected *p*_*cis*_ ∈ {2%, 5%} SNPs in cell type 2 as cis-SNPs, with their true effects following

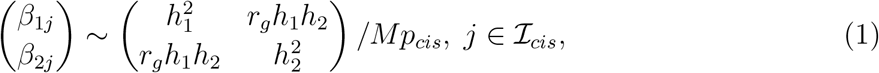

where *ℐ*_*cis*_ collected all indices of cis-SNPs, and the effects of the remaining SNPs were set to 0. We set cis-heritability *h*_1_ = 0, *h*_2_ ∈ {1%, 5%, 10%} and genetic correlation *r*_*g*_ = 0. We generated gene expression levels for both cell types according to

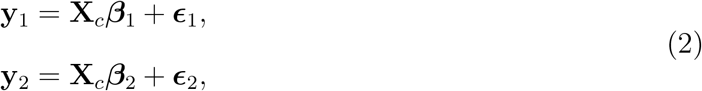

where ***ϵ***_1_ and ***ϵ***_2_ are the independent error terms. To generate tissue-level gene expression, we first generated the individual proportions of cell type 1 in the tissue samples from a beta distribution *π*_1*i*_ ∼ Beta(*απ*_1_, *α*(1 − *π*_1_)), *i* = 1, …, *N*_*t*_, where *π*_1_ ∈ {0.01, 0.05, 0.2, 0.5, 0.8, 0.95, 0.99} is the true mean proportion of cell type 1 and *N*_*t*_ is the tissue sample size, then the proportions of cell type 2 is given as 1 − *π*_1*i*_, and we set *α* = 50. Finally, we generated gene expression levels for the tissue samples based on

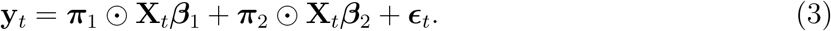

Four methods on eQTL analysis of cell type 1 were performed: only using data of cell type 1 (“Cell type 1”), using data of cell type 1 and 2 (denoted as “Cell type 1&2”), ieQTL analysis using tissue data (“Tissue ieQTL”), and IBSEP. We reported the fraction of SNPs with a *p*-value less than 0.05 for cell type 1 as the type I error rate. One can see from Fig. 2b that all the four methods produced well-controlled type I error rate regardless of heritability of cell type 2, fraction of cis-SNPs, as well as cell type proportions.

**Figure 2.**
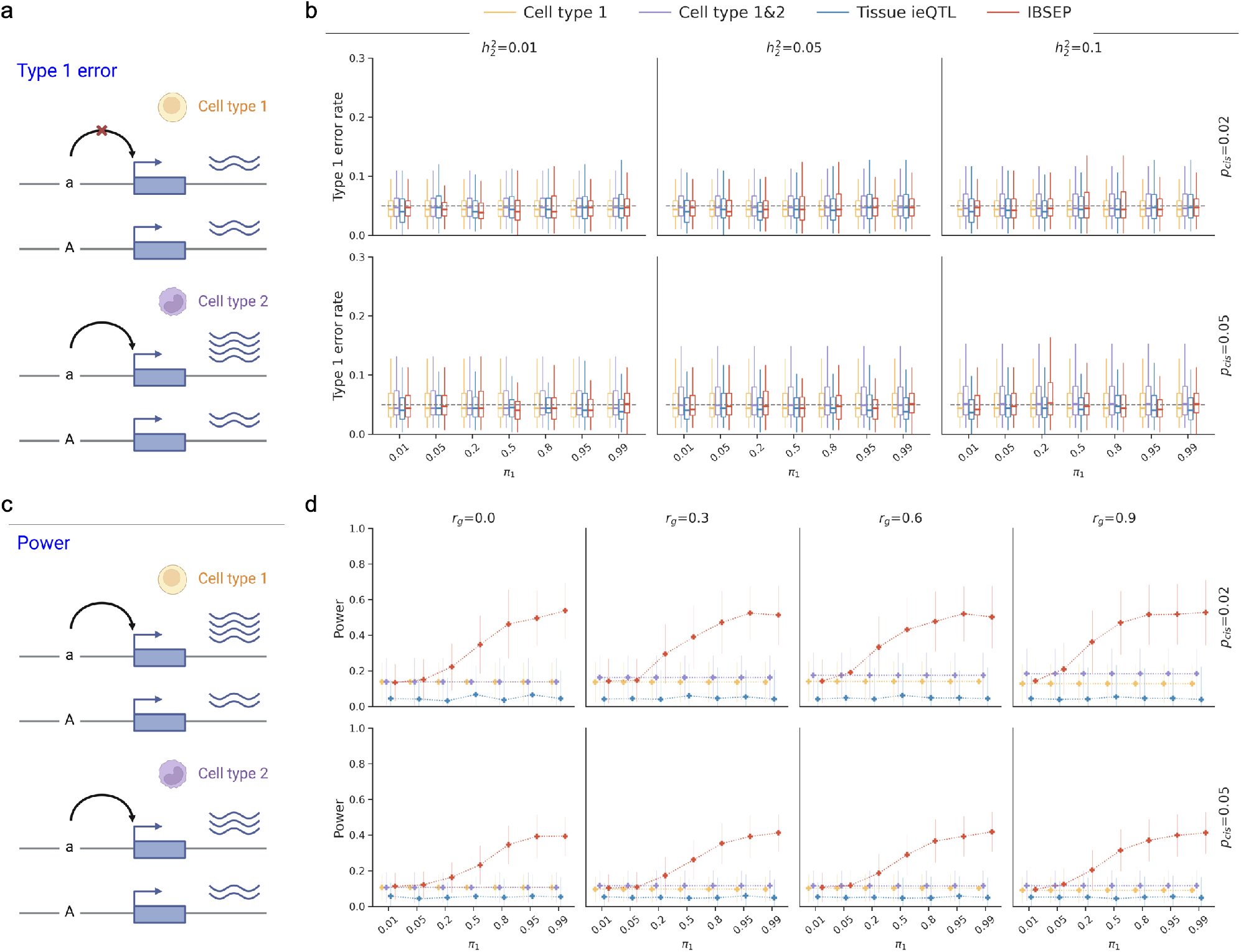
Simulation result. **a**, Diagram of simulation on type 1 error rate assessment. **b**, Type 1 error rates of IBSEP and compared methods under different heritability of cell type 2 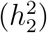, fractions of cis-SNPs (*p*_*cis*_) and mean proportions of cell type 1 in tissue samples (*π*_1_). **c**, Diagram of simulation on power assessment. **d**, Statistical power of IBSEP and compared methods under different genetic correlations between the two cell types (*r*_*g*_), fractions of cis-SNPs (*p*_*cis*_) and mean proportions of cell type 1 in tissue samples (*π*_1_).

Moreover, we conducted additional experiments to test the robustness of IBSEP under model mis-specification and inaccurate parameter estimation. In reality, there may be some other cell types present in tissue samples, that are either unknown or not available in single-cell dataset. Building upon the aforementioned simulation on type I error, let us assume the existence of cell type 3 in tissue samples, which is not available in the summary statistics of single-cell data. Nevertheless, IBSEP with the incorrect “2-cell-type” assumption can still control the type I error rate as long as the proportion of cell type 3 was not substantial. As expected, IBSEP with the correct “3-cell-type” assumption can control the type I error rate when single-cell data for cell type 3 was available (Fig. S4). Another realistic situation is when the cell type deconvolution method inaccurately estimate individual cell proportions in tissue samples, leading to biases between the estimated and true mean cell type proportion. We simulated various scenarios of inaccurate average cell type estimation and found that the bias in mean cell type proportion estimation has little impact on IBSEP’s type I error control (Fig. S5).

Next, we evaluated the power of IBSEP and compared methods. The procedure of generating genotypes and gene expressions are the same as above. We assumed that cell type 1 and cell type 2 shared cis-SNPs with their effect sizes also followed Eq. (1). Unlike the null simulation, we fixed 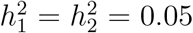, while varied genetic correlation between two cell types *r*_*g*_ ∈ {0, 0.3, 0.6, 0.9}. The power was calculated as the fraction of cis-SNPs with a *p*-value less than 0.05. As shown in Fig. 2d, IBSEP was the overall winner compared with other methods. Across different fractions of cis-SNPs and genetic correlations, the statistical power of IBSEP increased with the proportion of cell type 1 in the tissue samples. This is because as the proportion of cell type 1 in the tissue samples increases, IBSEP borrows more information from the tissue samples, leading to a more pronounced improvement in statistical power compared to the baseline model (“Cell type 1”). “Cell type 1&2” leveraged the genetic correlation between the two cell types, resulting in slight improvement over the baseline model and the degree of improvement became more significant with higher *r*_*g*_. However, when cell type 1 comprised a substantial proportion of the tissue samples (greater than 5%), IBSEP exceeded “Cell type 1&2”. This indicates that large-scale tissue samples can greatly assist in ct-eQTL prioritization. Lastly, we observed that the statistical power of “Tissue ieQTL” was notably lower than the baseline model, especially when both the heritability and cell proportion variance are small which is a typical scenario in practice. Although we can observed an improvement in the statistical power of “Tissue ieQTL” when 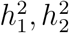 approached 1 and the variance of cell proportions was large, it still remained far below the baseline model S6. This suggests that conducting ct-eQTL analysis solely based on tissue samples may not achieve desirable outcomes.

### Real data analysis

We applied IBSEP for ct-eQTL mapping and downstream analysis in two tissues: blood and brain cortex. For both tissues, we obtained tissue-level eQTLs from GTEx and cell-type-level eQTLs, and integrated them using IBSEP to obtain improved ct-eQTL results. Compared to the original cell-type-level eQTL results, IBSEP ct-eQTLs showed significant improvements in statistical power. Specifically, IBSEP identified more ct-eQTLs, some of which have confirmed biological evidence, and some newly discovered ct-eQTLs were replicated in larger cell-type-level eQTL datasets. Furthermore, colocalization analysis between IBSEP ct-eQTLs and disease risk variants revealed more genes that potentially mediate disease development, further indicating that IBSEP has detected more eQTLs relevant to disease mechanisms in the cell type level.

### IBSEP in blood ct-eQTL analysis

#### Blood ct-eQTL prioritization

Due to the close association between gene expression in blood cells and many diseases, and the convenience of obtaining blood samples, there are rich data resources on tissue-level eQTL and cell-type level eQTL studies in blood. The analysis below involves two cell-type-level eQTL datasets: Oelen et al. sequenced and analyzed cell-type level eQTLs dataset in approximately 1 million PBMCs from 120 individuals (denoted as 1M sc-blood dataset), from which we obtained summary statistics for six cell types, including B cells, CD4 T cells, CD8 T cells, monocytes, natural killer (NK) cells, and dendritic cells (DC) [18]; the OneK1K eQTL dataset by Yazar et al. was derived from the analysis of scRNA-seq data from 982 individuals, covering 14 PBMC sub-cell types (which we grouped into the aforementioned six cell types) [17]. Additionally, we collected eQTL result for whole blood tissue from the GTEx project, which was obtained from the analysis of 670 samples with genotype information [4]. We performed integrative analysis of the 1M and GTEx datasets, while the OneK1K dataset, with its larger sample size, served as a replication dataset to validate the IBSEP result. Firstly, using xCell [19], we estimated the proportions of the six cell types in each GTEx blood sample, and then calculated the average cell type proportions. Subsequently, we input the cell-type-level eQTL summary statistics from 1M, tissue-level eQTL summary statistics from GTEx, and average cell type proportions into IBSEP to obtain the improved ct-eQTL summary statistics (Fig. 3a). We conducted a comprehensive and detailed comparison of the ct-eQTL results from 1M, OneK1K, IBSEP, and the tissue-level eQTL results from GTEx.

**Figure 3.**
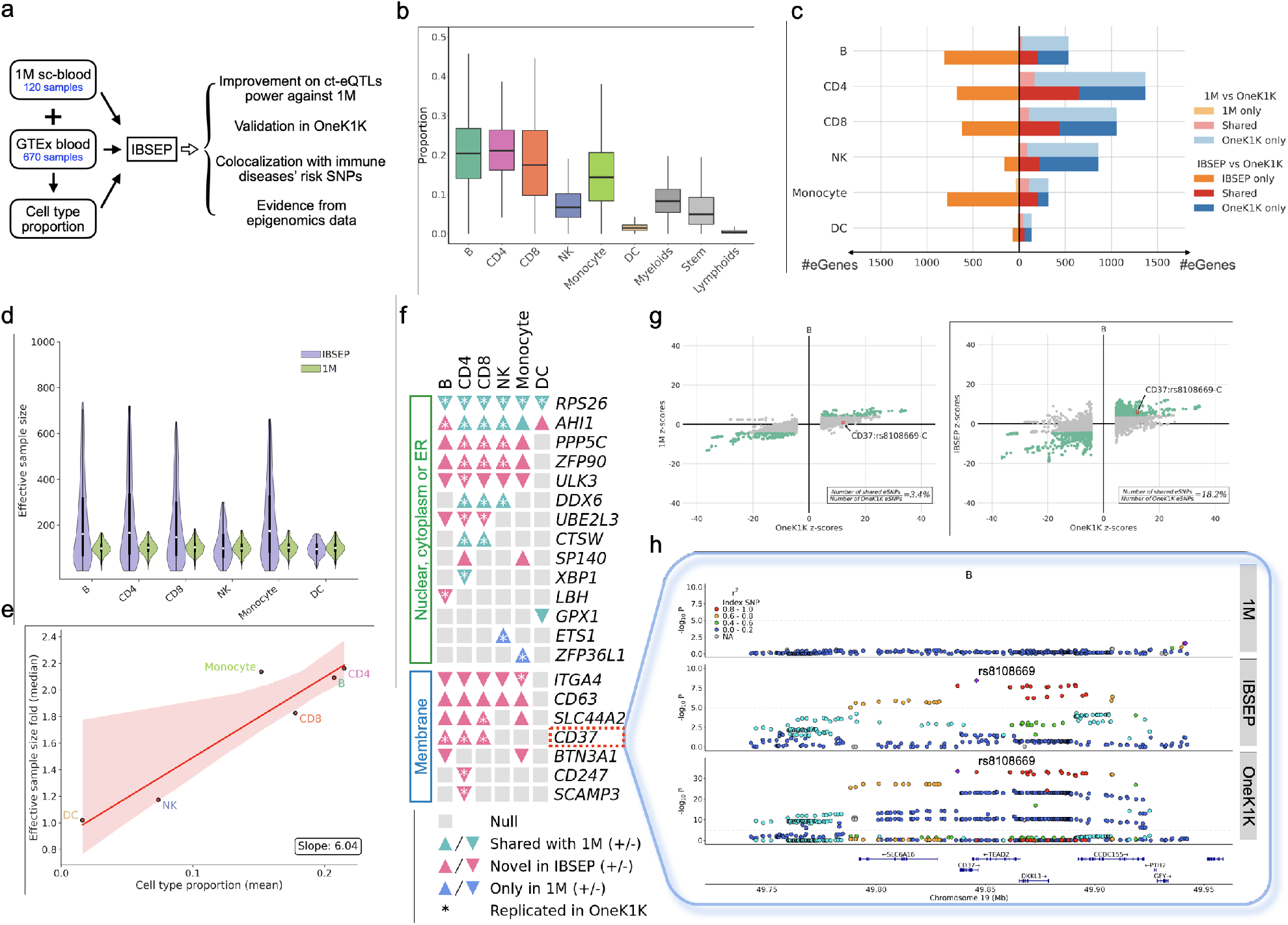
Results of blood ct-eQTL prioritization. **a**, Workflow of IBSEP’s blood ct-eQTL analysis. **b**, Boxplot of GTEx whole blood samples’ cell type proportions estimated by xCell. Cell types in xCell that are not among the six studied cell types are categorized into myeloid, stem, and lymphoid cells. **c**, Comparison of the number of eGenes discovered by 1M and IBSEP respectively with OneK1K in each cell type. **d**, Distribution of effective sample size of 1M and IBSEP for each gene in each cell type. **e**, Relationship between the average proportion of each cell type and the corresponding median effective sample size. **f**, ct-eQTLs of genes related to nuclei, cytoplasm or ER and membrane encoding discovered by 1M and IBSEP. Up/Down triangle represents the up/down-regulatory function of the effect allele of the lead ct-eQTL. **g**, Left: comparison of *z*-scores between OneK1K eSNPs and the same SNPs in 1M; Right: comparison of *z*-scores between OneK1K eSNPs and the same SNPs in IBSEP. **h**, Locuszooms of 1M, IBSEP and OneK1K around *CD37*.

As shown in Fig. 3b and Fig. S7, although the whole blood samples from GTEx contains cell types other than PBMCs, the combined average proportion of the six cell types of interest reaches 84%, with relatively low proportions of other cell types, which aligns with IBSEP’s requirement of a relatively low proportion of unobserved cell types. Among the studied six cell types, B cells, CD4 cells, CD8 cells, and monocytes have relatively higher average proportions, all exceeding 15% with the highest CD4 cells accounting for 21% on average; NK cells and DC have relatively low average proportions, at 7.4% and 1.6%, respectively. SNPs with a *p*-value less than 10^−5^ are defined as eSNPs, and genes containing at least one eSNP are referred to as eGenes. Fig. 3c shows the comparison of eGenes identified by 1M, OneK1K and IBSEP. First and foremost, it’s evident that IBSEP detects significantly more eGenes than 1M in all cell types. The increase in eGene count is positively correlated with the average cell type proportion of GTEx samples. For instance, CD4 cells and CD8 cells, which on average constitute 20% each in GTEx samples, exhibit approximately 6.5-fold and 8.3-fold increases in eGenes found by IBSEP, respectively. Conversely, NK cells and DC, which have lower average proportions in GTEx samples, show around 2.6-fold and 1.1-fold increases in eGenes found by IBSEP. Secondly, 70%-97% of eGenes identified by 1M are shared with OneK1K across the six cell types. Since the ct-eQTL analyses of both 1M and OneK1K were directly conducted using scRNA-seq data, the high consistency between the two datasets is reasonable. Hence, OneK1K to some extent reflects the ct-eQTLs to be discovered as sample size increases, making it a reference replication dataset to validate the credibility of IBSEP results. Despite a high fraction of eGenes identified by 1M overlap with OneK1K, 1M can only capture a small fraction of OneK1K eGenes, ranging from 5.6% in B cells to 32.8% in monocytes. In contrast, IBSEP identifies a larger number of OneK1K eGenes, ranging from 37.5% in B cells to 65.3% in monocytes. For instance, compared to 104 eGenes of NK cells identified in 1M datasets, IBSEP achieved a substantially higher power by identifying 378 eGenes, of which 57.7% can be replicated in OneK1K dataset (Fig. 3c). From another perspective, for each cell type, we compared the *z*-scores of 1M and IBSEP with OneK1K separately. It can be observed that, compared to 1M, IBSEP found more eSNPs in OneK1K (Fig. 3g, Fig. S10). For instance, in B cell, 1M only identified 3.4% of OneK1K eSNPs while IBSEP increased this proportion to 18.2%. At the same time, IBSEP and OneK1K show a high degree of consistency in the direction of allelic effects on OneK1K eSNPs, from 95.0% to 99.4%. In addition, IBSEP identifies some eGenes not found in OneK1K, most of which are included in GTEx (Fig. S12), which can be attributed to the tissue-level high-quality GTEx sources. Most importantly, although the sample size of GTEx is almost six times of 1M, IBSEP’s results are not dominated by GTEx as evidenced by the significantly higher proportion of IBSEP eGenes replicated in OneK1K than GTEx across all six cell types (Fig. S13).

When examining how many cell types each eGene is detected in, we found that 60% of 1M eGenes were only detected in one cell type, whereas this proportion decreased to 38% in OneK1K. This is because, as sample size increases, some eGenes become detectable in more cell types, which is consistent with the claim in the OneK1K paper [17]. Notice that despite utilizing GTEx data whose eQTLs are not in the cell type resolution, 32% of the IBSEP eGenes are present in only one cell type, a proportion higher than those of eGenes prioritized in other number of cell types (Fig. S14). These results demonstrate that IBSEP can simultaneously utilize both cell-type-level and tissue-level eQTL data, improving the resolution of many eGenes discovered at the tissue level to the cell type level. To quantitatively compare the gains in power, we calculated the effective sample sizes of both 1M and IBSEP for each cell type and each gene separately using the *χ*^2^ statistics (Fig. 3d). Overall, the median effective sample size of IBSEP is 1.06 to 3 times that of 1M. In other words, the sample size of 1M would need to increase by 6% to 200% to achieve an equivalent power gained by IBSEP. The improvement in statistical power of IBSEP primarily stems from tissue-level eQTLs with larger sample sizes: IBSEP is capable of extracting information proportional to cell type proportions from tissue-level eQTLs (Fig. 3e), integrating cell-type-level eQTLs, while also considering inter-cellular correlations and mitigating the influence of confounders, thus prioritizing more potential ct-eQTLs.

The ultimate goal of identifying ct-eQTLs is to establish links between genes and diseases through SNPs, thereby revealing cellular-level disease regulatory mechanisms. We focused on genes discussed in the OneK1K paper involved in two pathways: nuclei, cytoplasm or endoplasmic reticulum (ER) proteins encoding, and membrane proteins encoding, which are closely related to autoimmune diseases. As shown in Fig. 3f, due to its small sample size, 1M only identified 8 eGenes associated with nuclei, cytoplasm or ER and none for membrane. In contrast, IBSEP identified 19 eGenes, most of which can be replicated in OneK1K, although the corresponding cell types may differ. As a concrete example, *CD37* gene encodes the CD37 protein, a cell surface protein primarily expressed on B cells and T cells. Very consistently, IBSEP indeed newly identified *CD37* as an eGene for B cells, CD4 cells, and CD8 cells. From an ct-eQTL perspective, according to previous studies [20, 21], *CD37* is closely associated with B cells, involved in B cell adhesion and migration, and influences B cell activation, proliferation, and differentiation. In the locuszoom of *CD37* in B cells, rs8108669, initially not significant in 1M, became the most significant SNP affecting *CD37* expression in IBSEP, with its low *p*-value also observed in OneK1K. Furthermore, as shown in Fig. 3g, both IBSEP and OneK1K demonstrate that rs8108669-C is associated with upregulation of *CD37* expression, indicating the reproducibility of IBSEP. Besides B cells, rs8108669-C also displays salient transcriptional regulatory function of *CD37* expression in CD4 and CD8 cells, consistent with previous studies showing that *CD37* regulates T cell proliferation [22, 23]. From autoimmune diseases GWAS perspective, rs8108669-C has been confirmed to be associated with an increased risk of SLE [24]. The gene behind rs8108669-C, *CD37*, is considered an antigen and may play an important role in immune responses [25, 26]. Studies have found that the expression level of *CD37* may become abnormal in patients with autoimmune diseases such as systemic lupus erythematosus (SLE), multiple sclerosis (MS) and rheumatoid arthritis (RA) [27, 28, 29, 30]. These evidences clearly indicate that *CD37* play a critical role in the mechanism of autoimmune diseases through both B and T cells.

### Colocalization between blood ct-eQTLs and genetic risk SNPs of immune diseases

Next, we applied *Coloc* [31] to conduct colocalization analysis between ct-eQTLs and risk variants of four common autoimmune diseases, including RA [32], MS [30], IBD [33], and asthma [34]. *Coloc* is a Bayesian method which compares a gene’s eQTL *z*-scores and the corresponding GWAS *z*-scores to provide the posterior probability that the gene and the disease share a same causal SNP. We defined colocalized genes as those with the posterior probability greater than 0.7. Not surprisingly, for each disease and each cell type, more colocalized genes are identified using IBSEP’s ct-eQTLs than 1M (Fig. 4a). This can be attributed to IBSEP’s advantage in the power of ct-eQTLs prioritization compared to 1M, with a higher *z*-score for the true causal SNP, leading to an increased posterior probability of sharing the causal SNP. Taking RA for instance, CD4 cells play a central role in the pathogenesis of RA by orchestrating inflammatory responses and producing pro-inflammatory cytokines such as tumor necrosis factor-alpha (TNF-*α*) and interleukin-6 (IL-6) [35]. IBSEP identified the highest number of genes colocalized with RA in CD4 cells, highlighting the importance of CD4 cells in the mechanism of RA.

**Figure 4.**
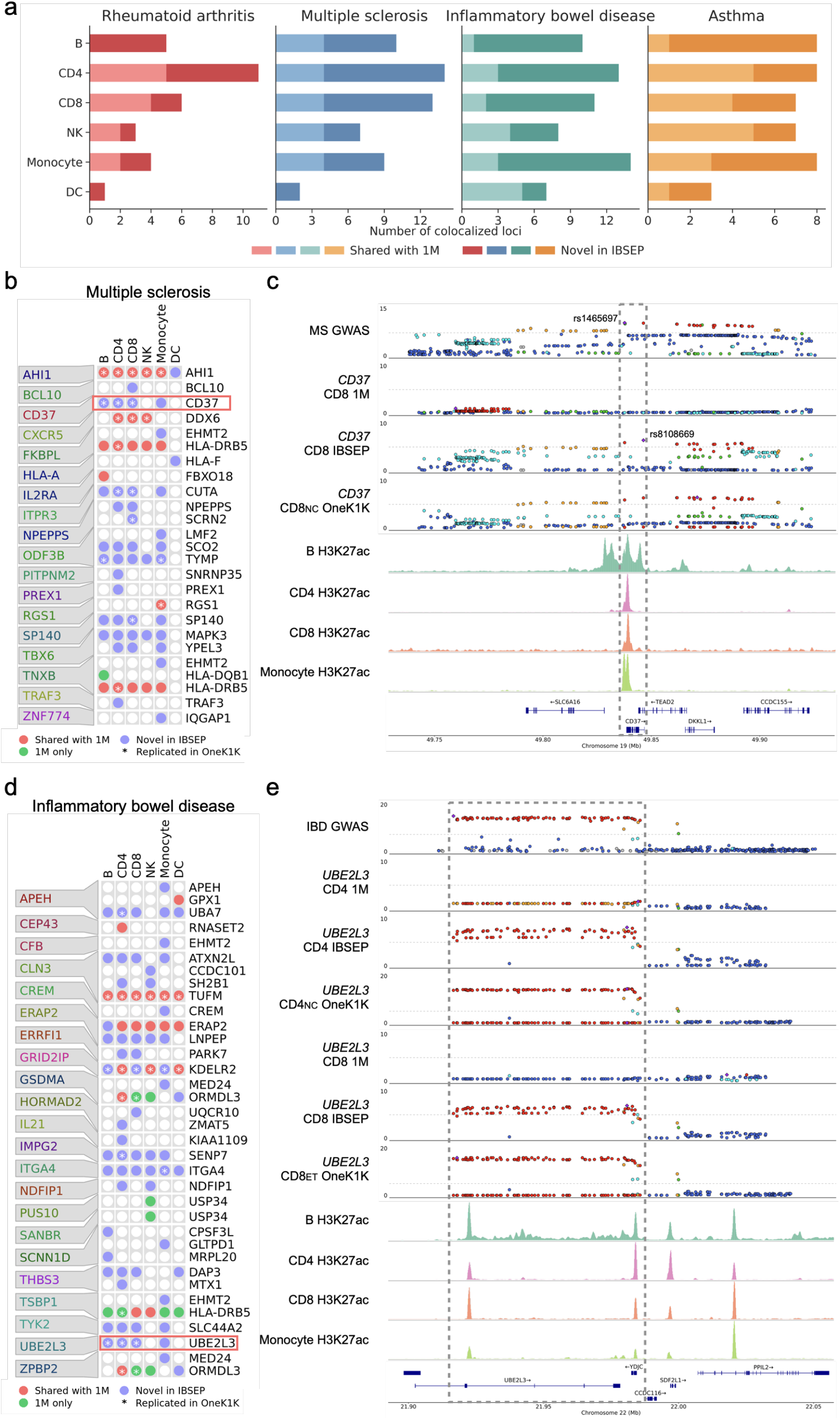
Results of colocalization between blood ct-eQTLs and genetic risk SNPs of immune disease. **a**, Comparison of the number of colocalized genes identified by 1M and IBSEP across four autoimmune diseases and six blood cell types. **b**, Cell-type-level colocalized genes of multiple sclerosis found by 1M and IBSEP. **c**, Locuszooms of GWAS, 1M, IBSEP and OneK1K around *CD37*, and corresponding H3K27ac tracks. **d**, Cell-type-level colocalized genes of inflammatory bowel disease found by 1M and IBSEP. **e**, Locuszooms of GWAS, 1M, IBSEP and OneK1K around *UBE2L3*, and corresponding H3K27ac tracks. In the locuszooms of GWAS and eQTL, the dashed lines represent -log *p*-value of 8 and 5, respectively.

For MS, IBSEP identified a total of 18 colocalized loci, containing 22 colocalized genes. Among them, 11 genes were displayed in only one cell type, 7 genes were displayed in 2-4 cell types, indicating a high cell type specificity of colocalized genes for MS (Fig. 4b). The aforementioned *CD37* gene, closely related to autoimmune diseases, was colocalized by IBSEP in four cell types: B cells, CD4 cells, CD8 cells, and monocytes. Particularly, in the first three cell types, the posterior probability of colocalization exceeded 0.98. As shown in Fig. 4c, the lead SNP for MS at the *CD37* locus was rs1465697, and the most significant SNP identified by IBSEP in CD8 cells was rs8108669, which is located near rs1465697 with a LD as high as 0.974. Moreover, the H3K27ac ChIP-seq tracks of B cells, CD4 cells, CD8 cells, and monocytes all showed peak signals at the position of rs1465697, corresponding to the promoter region of the *CD37* gene. The colocalization analysis and epigenomic evidence suggest that the association of the *CD37* gene with MS may be closely related to these four cell types.

For IBD, aberrant activation of the immune system is considered one of its key mechanisms, and multiple immune cell types are closely related to the pathogenesis of IBD. Combining the colocalization results of 1M and IBSEP, 22 IBD colocalized loci were identified in total, encompassing 31 colocalized genes (Fig. 4d). Among them, only one colocalized gene was exclusively discovered in 1M, and the vast majority of colocalized genes were newly identified by IBSEP. Notably, 14 genes were only expressed in a single cell type, while 13 genes were expressed in 4-6 cell types. This indicates that there are both cell-type-specific genes and genes with ubiquitous effects across multiple cell types relate to IBD, reflecting the complexity of the pathogenic mechanisms of IBD. As a specific example, IBSEP identified *UBE2LE* as a colocalized gene in B cells, CD4 cells, CD8 cells, and monocytes. *UBE2LE* is involved in regulating the activation, proliferation, differentiation, and cytokine production of immune cells, playing a crucial role in immune responses [36, 37]. Fig. 4e shows the locuszoom of GWAS and ct-eQTLs near the *UBE2LE* gene, as well as the corresponding H3K27ac ChIP-seq tracks. It is evident that GWAS, IBSEP, and OneK1K all exhibit consistently significant SNP signals along a high LD region of about 65kilobase (kb) from the start site of *UBE2LE* to its downstream, with significant peaks of H3K27ac at both ends, possibly corresponding to the promoter and enhancer of *UBE2LE*. These pieces of evidence suggest that *UBE2LE* may exert its influence on IBD through multiple cell types.

### IBSEP in brain ct-eQTL analysis

#### Brain ct-eQTL prioritization

In addition to blood tissues, brain cell-type-level eQTL analysis is another hot research topic. It is well known that the human brain is a highly complex organ, containing billions of neurons and various types of glial cells. Therefore, the mechanisms of brain disorders are complex, involving multiple cell types, each with its own distinct role, necessitating the study of brain ct-eQTLs. The largest existing dataset of brain single-cell level eQTLs is provided by Bryois et al. (the sc-brain dataset) [13], obtained by analyzing scRNA-seq from 192 independent individuals of 8 cell types, including excitatory neurons, inhibitory neurons, astrocytes, oligodendrocytes, microglia, oligodendrocyte progenitor cells (OPCs), endothelial, and pericytes. As the sample size of this dataset is not large enough, to identify more potential brain ct-eQTLs, we applied IBSEP to integratively analyze the results of the sc-brain dataset and the GTEx brain cortex eQTL dataset which contains 205 individual subjects, following a similar procedure as the blood tissue (Fig. 5a). First, we obtained the bulk RNA-seq data of GTEx brain cortex samples and estimated the proportions of the above-mentioned 8 cell types in each sample using Cibersortx [38] (Fig. 5b), and then applied the averages for integration (Fig. S17). Reasonably, neuronal cells (excitatory neurons and inhibitory neurons) account for approximately 41% on average, while glial cells (astrocytes, oligodendrocytes, microglia, and OPCs) account for approximately 51% on average, with only a small proportion of cells being endothelial and pericytes.

**Figure 5.**
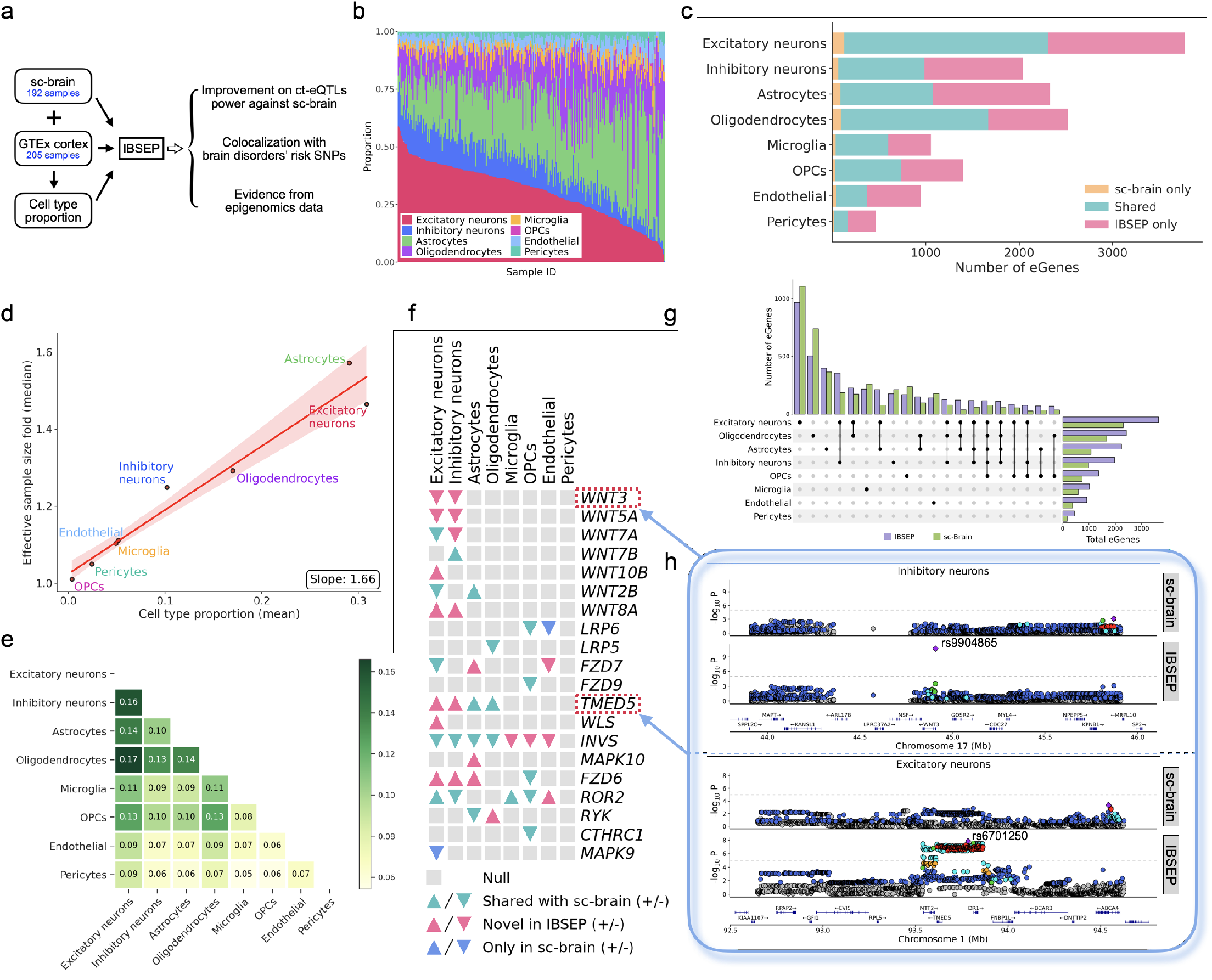
Results of brain ct-eQTL prioritization. **a**, Workflow of IBSEP’s brain ct-eQTL analysis. **b**, GTEx brain cortex samples’ individual cell type proportions estimated by Cibersortx. **c**, Comparison of the number of eGenes discovered by sc-brain and IBSEP in each cell type. **d**, Relationship between the average proportion of each cell type and the corresponding median effective sample size. **e**, Heatmap shown the fraction of genes with significant genetic correlation between cell types. **f**, ct-eQTLs of genes in Wnt pathway discovered by sc-brain and IBSEP. Up/Down triangle represents the up/down-regulatory function of the effect allele of the lead ct-eQTL. **g**, UpSet plot of the number of eGenes discovered by IBSEP and sc-brain across various cell types and their intersections. **h**, Locuszooms of sc-brain and IBSEP around *WNT3* and *TMED5*.

Fig. 5c compares the number of eGenes discovered by sc-brain and IBSEP across the eight cell types. Sc-brain identified 164-2307 eGenes, of which 89.6%-95.9% can be replicated by IBSEP. We observed that IBSEP maintained a high degree of consistency in allele effect direction with sc-brain on shared eSNPs (Fig. S20). In contrast, IBSEP achieved significant power gains by identifying 449-3644 eGenes, with a considerable proportion being newly discovered in each cell type. For instance, IBSEP identified 1464 newly discovered eGenes of excitatory neurons, which represented the largest proportion in GTEx samples and therefore more information could be leveraged from GTEx data; for microglia which represented a smaller proportion in GTEx samples, IBSEP identified 457 novel eGenes. Despite the sample size of GTEx is not significantly larger than sc-brain, IBSEP still demonstrates a substantial improvement in statistical power due to its high-quality RNA-seq data. In the analysis of blood tissues data, the majority of newly discovered eGenes by IBSEP overlapped with GTEx. However, in brain tissues, 36.4%-48.6% of newly discovered eGenes by IBSEP were not present in GTEx eGenes. To investigate how IBSEP discovered these novel eGenes, we conducted ct-eQTL analysis using only summary statistics from sc-brain (referred to as IBSEP-ct), meaning that IBSEP-ct only utilized genetic correlations between cell types without leveraging tissue-level information. Consistent with the simulation results, although the statistical power of IBSEP-ct was slightly lower than IBSEP, it still identified more eGenes compared to sc-brain (Fig. S21). This is because some cell types exhibit significant genetic correlations with other cell types for certain genes, such as excitatory neurons with inhibitory neurons, oligodendrocytes, and astrocytes, allowing IBSEP-ct effectively utilize this to improve statistical power (Fig. 5e). We found that 81%-96% of eGenes newly discovered by IBSEP were included in GTEx or IBSEP-ct (Fig. S19,S22), and the number of novel eGenes present exclusively in GTEx but not in IBSEP-ct was highly positively correlated with the average cell type proportion in GTEx samples (Fig. S23). These evidence indicate that the novel eGenes can be attributed to IBSEP leveraging both inter-cell type genetic correlations and tissue-level eQTLs.

When sorting the number of eGenes by cell type count, we observed that the ranks were highly consistent between IBSEP and sc-brain: the highest number of eGenes was observed in only one cell type, and the number decreases as eGenes were observed in more cell types (Fig. S24). However, it is evident that IBSEP, compared to sc-brain, increased the proportion of eGenes observed in multiple cell types, which is a natural outcome of statistical power improvement[17]. More specifically, as shown in Fig. 5g, eGenes specific to excitatory neurons, oligodendrocytes, and astrocytes ranked among the top three most in both sc-brain and IBSEP, which is related to the abundance of these three cell types in scRNA-seq and GTEx samples. Among eGenes observed in two cell types, the top-ranking pairs include excitatory neurons-inhibitory neurons, excitatory neurons-oligodendrocytes, excitatory neurons-astrocytes, and oligodendrocytes-astrocytes, which aligns with their higher statistical power and biological correlation between these cell types. We further calculated the effective sample size of sc-brain and IBSEP in each cell type. As expected, the effective sample size of IBSEP was generally higher than that of sc-brain in all cell types (Fig. S25). Specifically, the median effective sample size of IBSEP was 1.28-1.74 times that of sc-brain, and these folds were significantly positively correlated with the average proportion of cell types in GTEx samples (Fig. 5d).

The development and function of the nervous system are regulated by multiple signaling pathways. The Wnt pathway is one of the important signaling pathways in neurons for maintaining the normal function and structure of the nervous system [39, 40]. Using a public human brain single cell atlas [41], we observed that neurons exhibit higher Wnt pathway gene expressions than other cell types (Fig. S26). Therefore, we focused on examining the ct-eQTLs of genes related to the Wnt pathway and compared the results between IBSEP and sc-brain for these genes. As shown in Fig. 5f, IBSEP identified 19 eGenes associated with the Wnt pathway, 6 of which are novel compared to sc-brain. Most importantly, among these eGenes discovered by IBSEP, excitatory neurons or inhibitory neurons showed significant enrichment compared with other cell types, which was in line with crucial biological functions of Wnt pathway in neurons. Specifically, IBSEP identified 7 genes in *WNT* family, all of which were displayed in neurons. The *WNT* gene family is core to the Wnt pathway and is associated with various brain disorders [42, 43]. For example, *WNT3* is linked to several disorders including depression, Alzheimer’s disease, and Parkinson’s disease [44, 45, 46]. IBSEP found *WNT3* to be an eGene for excitatory neurons and inhibitory neurons, whereas sc-brain failed to identify it. As shown in the upper plot of Fig. 5h, the eQTL signals for *WNT3* in inhibitory neurons are weak in sc-brain. By contrast, IBSEP successfully identified a significant eQTL, rs9904865, in the promoter region of *WNT3*. Another example is *TMED5*, a gene reported to be associated with educational attainment and Alzheimer’s disease[47, 48]. sc-brain identified *TMED5* as an eGene for astrocytes and oligodendrocytes only, while IBSEP additionally found it to be an eGene in neurons. As shown in the lower plot of Fig. 5h, there was no significant signal for *TMED5* in sc-brain on excitatory neurons, whereas IBSEP showed strong signals around *TMED5* spanning a ∼300kb region. Besides neurons, the Wnt pathway also regulates the function and interaction of glial cells in the nervous system [49], and correspondingly, IBSEP discovered a number of eGenes displayed in glial cells.

#### Colocalization between brain ct-eQTLs and genetic risk SNPs of brain disorders

To explore the role of specific cell types in brain disorders, we conducted colocalization analysis for brain ct-eQTLs and risk loci from four brain disorders GWAS, including Alzheimer’s disease (AD) [50], bipolar disorder (BIP) [51], major depression disease (MDD) [52], and schizophrenia (SCZ) [53]. The power of colocalization analysis was strongly enhanced with IBSEP results: using sc-brain’s ct-eQTL results, we can only identify 24-120 colocalized loci across the four diseases, while leveraging IBSEP’s ct-eQTL results revealed 34-191 colocalized loci (Fig. 6a). Furthermore, for each disease, IBSEP maintained a generally consistent relative abundance of colocalized loci across cell types compared to sc-brain. For instance, regarding AD, both IBSEP and sc-brain identified most colocalized loci in microglia, in concordance with the accumulated scientific understanding of the critical role of microglia in AD [54]; for the other three psychiatric disorders, both IBSEP and sc-brain found more colocalized loci in neurons.

**Figure 6.**
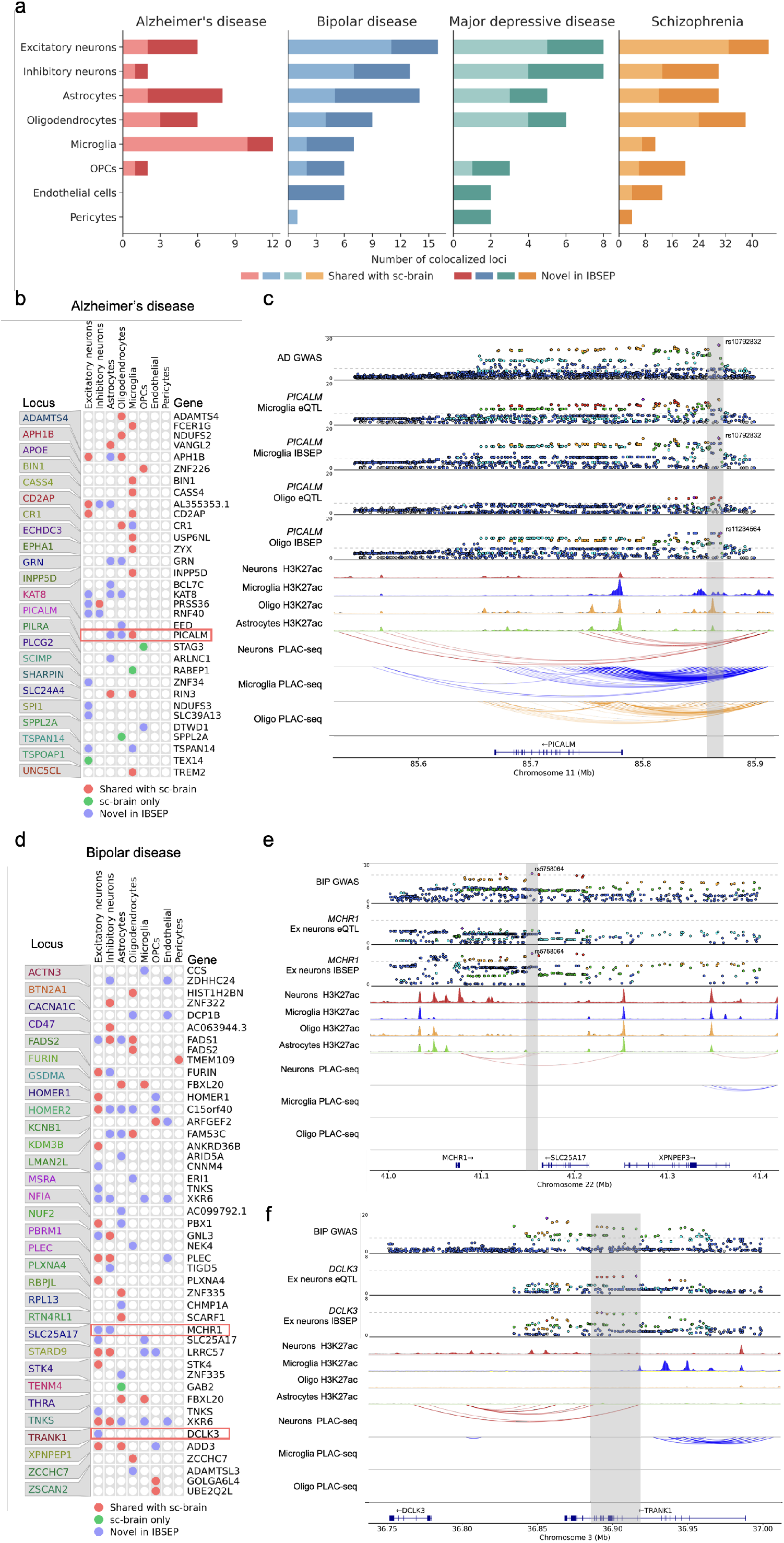
Results of colocalization between blood ct-eQTLs and genetic risk SNPs of brain disorders. **a**, Comparison of the number of colocalized genes identified by sc-brain and IBSEP across four brain disorders and eight brain cell types. **b**, Cell-type-level colocalized genes of Alzheimer’s disease found by sc-brain and IBSEP. **c**, Locuszooms of GWAS, sc-brain and IBSEP around *PICALM*, and corresponding epigenomic tracks. **d**, Cell-type-level colocalized genes of bipolar disease found by sc-brain and IBSEP. **e**, Locuszooms of GWAS, sc-brain and IBSEP around *MCHR1*, and corresponding epigenomic tracks. **f**, Locuszooms of GWAS, sc-brain and IBSEP around *DCLK3*, and corresponding epigenomic tracks. In the locuszooms of GWAS and eQTL, the dashed lines represent -log *p*-value of 8 and 5, respectively.

As a concrete example, sc-brain and IBSEP identified 23 and 29 colocalized genes in AD, respectively, which exhibited a high degree of cell type specificity. Among the colocalized genes identified by IBSEP, 18 genes were exclusively expressed in one cell type, while 7 genes and 4 genes were present in 2 and 3 cell types, respectively. Microglia plays a crucial role in clearing amyloid-beta (A*β*) plaques, abnormal aggregation of which is a major pathological feature of AD. IBSEP identified several genes that are colocalized exclusively in microglia. For instance, *BIN1* has been found to be a significant risk locus for AD [55], with its expression levels influenced by a microglia-specific enhancer [56]. Another microglia-specific colocalized gene shared by IBSEP and sc-brain was *INPP5D*, whose expression levels positively correlate with amyloid plaque density and increase predominantly in plaque-associated microglia with the progression of AD [57]. In addition to microglia, AD is also associated with other cell types. As mentioned earlier, IBSEP has discovered some genes colocalized in two or more cell types. For instance, astrocytes play crucial roles in neuronal metabolism and clearance of A*β*, and the development of astrocytes with pathological phenotypes may contribute to the progression of AD. Both IBSEP and sc-brain have colocalized *RIN3* in astrocytes and microglia, which has been reported as a potential causal gene for AD [58]. Previous studies have found that *RIN3* plays a significant role in the early endocytic pathway, which is closely related to microglial function [59]. Although the specific interactions and functions between *RIN3* and astrocytes have not been fully elucidated, cell-type-specific eQTLs detected by ISBEP may shed light on the genetic etiology of AD. Another frequently discussed gene, *PICALM*, has been reported to be highly expressed in microglia in late-onset AD [60]. Both sc-brain and IBSEP identified *PICALM* colocalized in microglia. Additionally, IBSEP found *PICALM* colocalized in oligodendrocytes and astrocytes as well. In Fig. 6c, significant GWAS and eQTL signals were observed at ∼80kb upstream of *PICALM*, with clear peaks in H3K27ac ChIP-seq in microglia as well as oligodendrocytes and astrocytes. PLAC-seq in both microglia and oligodendrocytes shows interaction with the gene’s starting position in this region, suggesting it may be an active enhancer for *PICALM* in multiple cell types rather than microglia specific. On the contrary, there are no significant epigenomic signals in neurons in this region, consistent with both IBSEP and sc-brain not colocalizing *PICALM* in neurons.

For BIP, sc-brain identified 27 colocalized genes, while IBSEP increased this number to 41. The pathogenesis of BIP is complex, involving multiple cell types, particularly neurons and glial cells, with the former’s abnormal activity potentially leading to mood fluctuations and the latter playing a crucial role in maintaining neuronal function and regulating neurotransmitter levels [61, 62, 63]. Among the 41 colocalized genes identified by IBSEP, 27 were observed in excitatory or inhibitory neurons, and 19 were associated with astrocytes or oligodendrocytes, highlighting the importance of these cell types in the pathogenesis of BIP. Mullins et al. conducted causal analysis using the same GWAS summary statistics with the tissue-level brain eQTLs from PsychENCODE Consortium [64], identifying several genes potentially mediating BIP [51]. Some of these genes were also colocalized by both IBSEP and sc-brain, including *GNL3, ADD3, FURIN, LRRC57*, and *STK4*. Most importantly, IBSEP can identify the cell types in which these genes act as causal mediators, enabling us to uncover biological pathogenic mechanisms at a fine-grained scale. For instance, *FURIN* is reported to be associated with multiple psychiatric disorders [53, 65, 66]. The most significant SNP of *FURIN*, rs4702, was identified by BIP GWAS as well as IBSEP in neurons. In addition, IBSEP further colocalized *MCHR1* and *DCLK3*, which are also in the list of potential causal genes [51] but not discovered by sc-brain. The *MCHR1* gene, possibly involved in mood regulation and circadian rhythm disruption, is associated with anxiety-like behaviors ([67, 68]). IBSEP identified it as a colocalized gene in both excitatory neurons and inhibitory neurons. As shown in Fig. 6e, the most significant SNP, rs5758064, of IBSEP in excitatory neurons is also the lead SNP for this locus in the BIP GWAS. Epigenetic signals indicate that rs5758064 may correspond to a neuron-specific enhancer interacting with *MCHR1. DCLK3* plays a critical role in the development and maintenance of the nervous system, which encodes a kinase regulating multiple signaling pathways within cells [69]. In the region of 36.88-36.92 Megabase (Mb) of chromosome 3, both excitatory neurons eQTL and BIP GWAS exhibited significant signals. The PLAC-seq shows interaction between *DCLK3* and this region, suggesting it may corresponds to a neuron-specific enhancer for *DCLK3*.

## Discussion

Ct-eQTLs mapping are critical for understanding the genetic regulation of gene expression in specific cellular contexts, which in turn informs the mechanisms underlying complex diseases. This paper introduces a novel framework, IBSEP, designed to enhance the detection and prioritization of ct-eQTLs by integrating bulk RNA-seq and scRNA-seq data. By leveraging the strengths of both data types, IBSEP overcomes the limitations of each, resulting in improved accuracy and detection power for ct-eQTLs. Through comprehensive simulations, we showed that our method well controls the type I error rate while having greater statistical power compared to existing approaches, and is robust across various genetic architecture settings. Our analyses of peripheral blood mononuclear cells and brain cortex datasets demonstrate that IBSEP greatly enhances the power of ct-eQTLs prioritization, uncovering new transcriptional regulatory mechanisms specific to different cell types. These findings provide valuable insights into the genetic architecture of complex traits at a cellular resolution.

Genetic variations can influence RNA levels and regulate various molecular traits, such as histone modifications, chromatin accessibility, allele-specific expression, alternative splicing, DNA methylation, and protein expression[70]. QTL analyses based on histone modifications and chromatin accessibility can reveal variant loci that affect transcription factor binding and nucleosome positioning; Detection of protein QTLs (pQTLs) can identify variant loci impacting transcriptional and post-transcriptional mechanisms. However, these molecular QTLs (xQTLs) studies face similar challenges to eQTL studies, with most remaining at the tissue level due to the high costs and technical constraints limiting cell type-specific QTL research. For instance, the recently released UK Biobank Pharma Proteomics Project includes 2,923 plasma protein traits from 54,219 samples [71, 72]. In contrast, a recent cell type-specific pQTL study had only 303 samples [73], resulting in many potential ct-pQTLs being undetected due to insufficient statistical power. As a general framework for integrating tissue-level and cell-type-level QTL data, the principles and methodologies of IBSEP can be extended to these molecular traits straightforwardly. By incorporating these additional layers of molecular data, researchers can develop a more comprehensive understanding of how genetic variants influence a wide array of biological processes. This expansion is crucial for capturing the full scope of gene regulation and its effects on phenotype and disease. The integration of diverse molecular traits with ct-eQTL analysis will provide a more detailed and nuanced view of genetic regulation.

Due to the genetic heterogeneity among different populations, QTLs identified in studies of genetic mechanisms of specific molecular traits can vary across populations [74, 75]. Unfortunately, most current eQTL samples come from European population. As many have noted, genomic discoveries in Europeans cannot be directly translated to non-European individuals [76], resulting in severe underrepresentation of non-European populations in eQTL research. This exacerbates inequalities in genetic research and healthcare for non-European populations. In fact, population-specific genetic variants and environmental contexts can significantly affect the manifestation of eQTLs. Studying diverse populations can reveal the different transcriptional regulatory patterns and different genetic etiology for diseases [77]. Therefore, in future research, we plan to extend the integrated analysis method applied in IBSEP to diverse ethnic groups. We aim to improve the statistical power of ct-eQTL localization in non-European populations and identify both shared and population specific ct-eQTLs, thereby improving the generalizability and applicability of findings and ensuring that medical insights and treatments are relevant and effective for a broad range of individuals.

Following eQTL detection, statistical fine-mapping can identify candidate causal variants likely to drive variation in expression within specific cell types, improving the translation of findings of genetic studies [78, 79]. Particularly, Mendelian Randomization (MR) is widely used to investigate the causal relationship between exposure factors and disease outcomes [80]. In the context of ct-eQTLs, the expression level of a gene in a specific cell type is considered the exposure factor, while the complex disease is regarded as the outcome. The candidate causal variants inferred from ct-eQTLs results could be used as instrumental variables, which typically come from a small region of the genome surrounding the target gene, known as cis-variants, and thereforely this type of analysis is referred to as cis-Mendelian Randomization (cis-MR) analysis. With such causal inference framework, cis-MR can use candidate causal variants to identify drug targets that increase disease risk through alterations in gene expression, enabling pinpoint causal variants with greater accuracy and understand the biological mechanisms linking genetic variants to disease in distinct cellular resolution.

Although IBSEP provides a effective way to leverage the advantages of each type of data to improve the statistical power for ct-eQTLs prioritization. We should be aware of some potential limitations. First, in the estimation of cell type proportions (***π***) for bulk RNA-seq samples, typically count scale gene expressions data were used. However, in the Bayesian hierarchical model applied by IBSEP (Eq. 5), we directly plugged in the ***π*** into the quantile normalized gene expressions models. This disparity arises from the different purposes and contexts in which ***π*** is utilized. The initial estimation of cell type proportions using count data allows for a robust and unbiased assessment of the relative abundance of each cell type. The purpose of incorporating these pre-estimated cell type proportions in IBSEP is merely to ensure that the model captures the relative differences of cell type compositions in tissue-level data more effectively. Besides, we note that even in the presence of substantial cell type proportion estimation errors, IBSEP still can well control type I error control rate (Fig. S5). Second, IBSEP can leverage the widespread genetic correlations among cell types, especially biologically close cell types, for certain genes to further boost statistical power (Fig. 5e). However, the estimation of co-heritability using LDSC regression [81] may be unstable due to the limited number of cis-SNPs in a gene. Therefore, we adopt a conservative approach by reducing the co-heritability estimates to zero if it do not significantly deviate from zero. Third, IBSEP is designed to identify ct-eQTLs across discrete cell types, such that the tissue-level gene expressions from bulk RNA-seq can be decomposed into weighted average of gene expression at the cell type level. Recently, a few single cell eQTL studies have begun to focus on continuous phenotypes or intermediate states instead of discrete lineages, aiming to map “dynamic” eQTLs that exhibit dynamical effects along a continuous axis. This strategy have been successfully applied to continuous trajectories within differentiating iPS cells [82] and T cells[83]. How to improve IBSEP model to accommodate these granular single-cell resolution data is an interesting direction for future work.

## Methods

### The IBSEP model

We begin our formulation with the individual-level scRNA-seq and bulk RNA-seq data. For the tissue level data of a target gene, suppose we have collected a dataset {**y**_*t*_, **X**_*t*_}, where 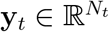 is the normalized gene expression vector and 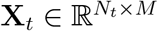 is the standardized genotype matrix. *M* is the number of SNPs within the target gene (e.g., 200) and *N*_*t*_ is the number of tissue samples (e.g., 1000). Without loss of generality, we assume the covariates have been properly adjusted. More detailed treatment on covariates adjustment can be found in our previous work [84, 85]. The following linear model relates gene expression and genotype:

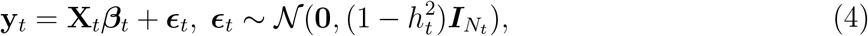

where 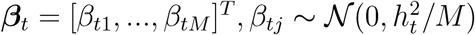 are the cis-SNP effect sizes, 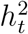 is the cis-heritability of the target gene and ***ϵ***_*t*_ is the independent error term.

For the sake of illustration, consider only two cell types in the tissue. Suppose by single cell sequencing, we have obtained a cell type level eQTL dataset {**y**_1_, **y**_2_, **X**_*c*_}, where 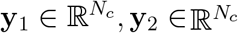 are normalized mean gene expression vectors of cell type 1 and cell type 2, respectively, 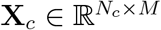 is standardized genotype matrix, and *N*_*c*_ is sample size of scRNA-seq data which is typically smaller than 200. Combining tissue level and cell type level data, we consider the Bayesian hierachical linear model:

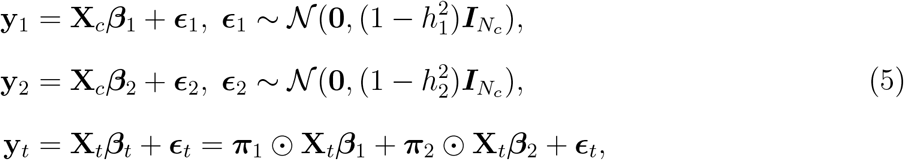

where 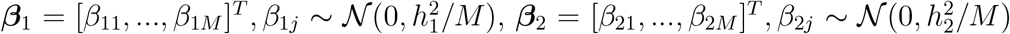 are the cis-SNP effect sizes of cell type 1 and cell type 2, respectively, 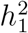 and 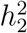 are the cis-heritability of this gene in the two cell types, ***ϵ***_1_ and ***ϵ***_2_ are the independent error terms, 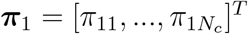 and 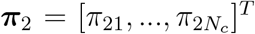 are proportions of the two cell types in the tissue samples, and ⊙ represents element-wise product. Thus, tissue-level gene expression **y**_*t*_ can be mathematically decomposed into a weighted average of cell type 1 (**X**_1_***β***_1_) and cell type 2 (**X**_2_***β***_2_) where the weights are the cell type proportions ***π***_1_ and ***π***_2_. We aimed to estimate the cis-SNP effect sizes (***β***_1_, ***β***_2_) by integrating tissue level and cell type level data. However, the variation in cell type proportions within tissue samples presents significant challenges for integrating these two sets of models. On the one hand, the need to account for the heterogeneity of samples makes it difficult to apply to summary level eQTLs data. On the other hand, it is challenging to design an efficient and reliable algorithm that can simultaneously analyzing multiple cell types with mutual influences in real ct-eQTLs analysis.

### The IBSEP model with summary-level data

To illustrate the challenges, assume the individual data {**X**_*t*_, **y**_*t*_, **X**_*c*_, **y**_1_, **y**_2_} are unavailable, but we have access to the summary statistics 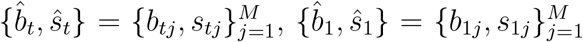 and 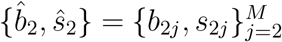:

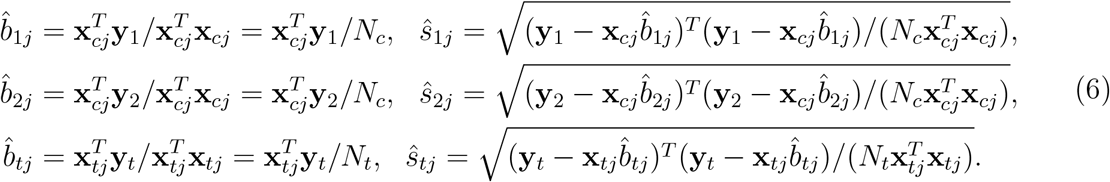

Clearly, the eQTLs and ct-eQTLs obtained through independent analyses fail to fully leverage the correlations between ***β***_1_, ***β***_2_, ***β***_*t*_, resulting in suboptimal statistical power. To bridge the tissue-level and cell-type-level eQTL data in mathematical models, we define the true marginal effect sizes as

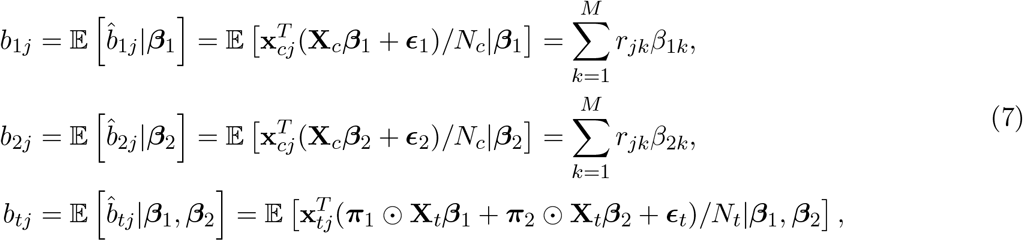

where *r*_*jk*_ is the correlation between SNP *j* and SNP *k*. In the last line of Eq. (7), it can be observed that the differences in ***π***_1_ and ***π***_2_ across different samples make it difficult to establish a direct relationship between the tissue-level marginal effects **b**_*t*_ and the cell-type-level marginal effects {**b**_1_, **b**_2_}.

Fortunately, we found that when integrating summary statistic level eQTL data, differences in cell type proportions in tissue samples can be disregarded. The cell type proportion vector ***π***_1_, ***π***_2_ of a given sample in tissue-level RNA sequencing data can be approximated by their mean 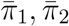, respectively. Specifically, we proved that when the sample size is large enough, the estimate of marginal effect size 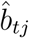 for cis-SNP *j* in a given tissue *t* is approximately independent of the cell type proportion composition variations across individuals. In other words, the estimation of 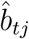 can be approximated by the mean cell type proportions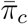’s instead of requiring individual *π*_*cj*_’s. The proof is provided in Supplementary Section “Theoretical justification on the usage of mean cell type proportions”, and the experimental validation is given latter. Given this point, we update Eq. (5) as follows:

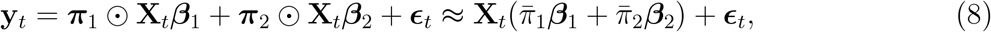

and the marginal effect *b*_*tj*_:

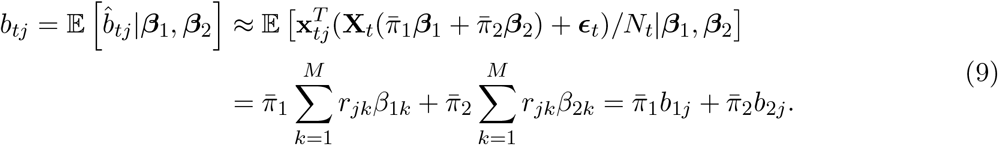

Based on this, we can finally establish a simple yet effective linear model to integrate tissue-level data and cell-type-level data.

In order to better utilize the correlation among ***β***_1_, ***β***_2_ and ***β***_*t*_, we impose the following probabilistic structure for ***β***_1_ and ***β***_2_:

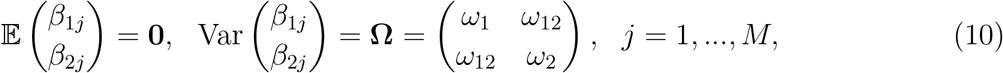

where **Ω** captures the genetic covariance of the two cell types in cis-SNPs. The diagonal elements represent the per-SNP cis-heritability for cell type 1 and 2, and the off-diagonal elements represent the per-SNP cis co-heritability between the two cell types.

The estimates of marginal effect sizes 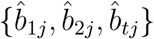 for cis-SNP *j* can be decomposed as

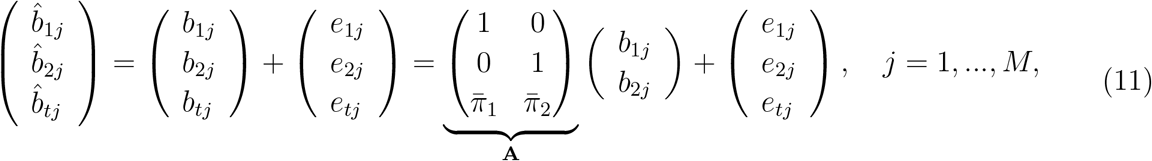

where *e*_1*j*_, *e*_2*j*_ and *e*_*tj*_ are the independent estimation errors with 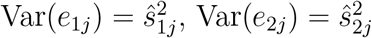 and 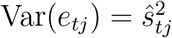.

With the model specification, we can derive the covariance of marginal effect sizes (Eq. S2-S6). In a compact form, we have

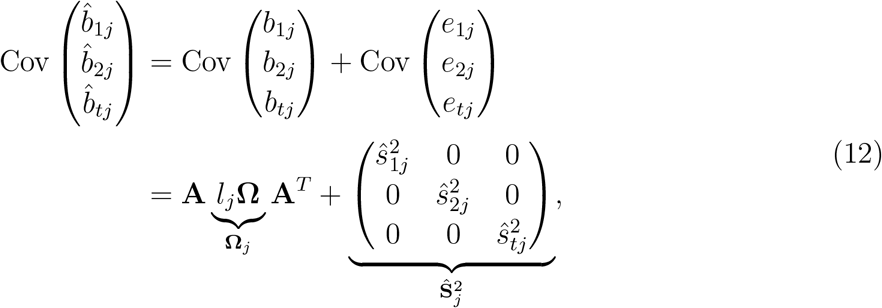

where 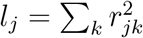 is LD scores for SNP *j*.

To account for the hidden confounding biases in GWAS summary statistics, we generalize the linear model (8) based on the genetic drift model used in LDSC [81] and obtain the modified covariance of marginal effects (Eq. S7). Now Eq. 12 becomes:

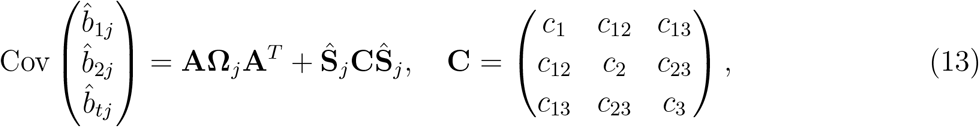

where elements in **C** are inflation constants that adjust for the confounding biases due to the geographic structure or sample overlap etc.

We summarize the reasons and benefits of the proposed model. First, by integrating tissue-level marginal effect *b*_*tj*_ and the cell-type-level marginal effect *b*_1*j*_, *b*_2*j*_ using a simple linear model, we can effectively combine high-quality large-scale bulk RNA-seq data with small-sample scRNA-seq data, fully leveraging the advantages of each type of data to enhance the statistical power of ct-eQTLs prioritization. Second, by directly modeling the eQTLs summary statistics, we can use these summary statistics as input, providing greater utility in large-scale data integration analyses. Third, by introducing off-diagonal elements Ω to represent the average cis-coheritability between SNPs across cell types, we can consider the complex structural relationships between cell types, revealing both shared and specific patterns of transcriptional regulation across different cell types.

### The IBSEP estimator

Based on the proposed statistical model for integrating ct-eQTLs analysis, the following focuses on the development of efficient algorithms tailored to it. Based on Eq. (11) and Eq. (13), we first obtain the conditional mean of the estimated marginal effects

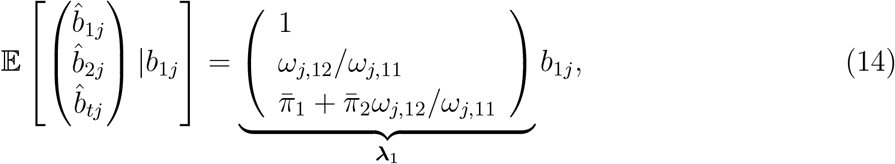

and the conditional variance

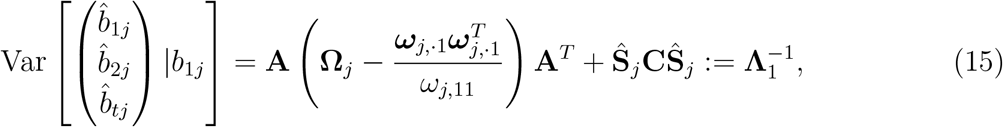

where ***ω***_*j,·*1_ = [***ω***_*j*,11_, ***ω***_*j*,12_]^*T*^ is the first column of **Ω**_*j*_. Based on Eq. (14), we can define 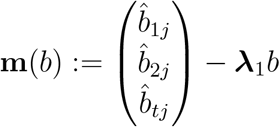 and give the first-order moment condition

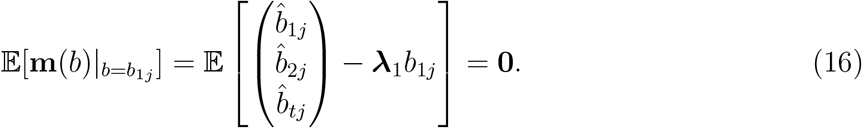

In the framework of Generalized Method of Moments (GMM) [86], we treat the marginal effect *b*_1*j*_ of cell type 1 as an estimable parameter and obtain its best linear unbiased estimator by solving the moment condition Eq. (16):

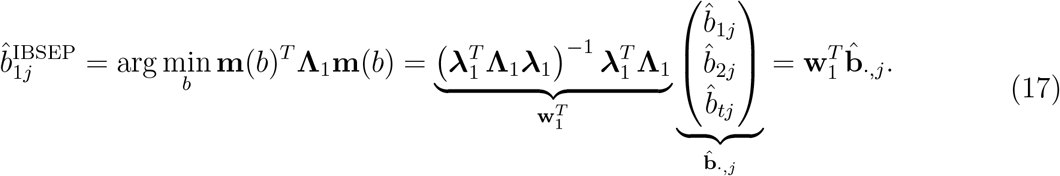

According to the asymptotic normality of GMM, its corresponding variance is

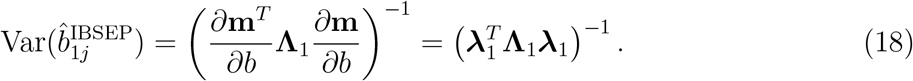

### Parameter estimation and ct-eQTL prioritization

The above IBSEP estimator involves a few unknown parameters to be estimated, including mean cell type proportions 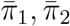, cis-heritability of the two cell types (*ω*_1_, *ω*_2_), the cis-coheritability between the two cell types (*ω*_12_) and inflation constants in **C**.

To estimate the mean cell type proportions, one can feed the bulk RNA-seq into a commonly used cell type deconvolution method such as xCell [19] and Cibersortx [38] to obtain individual cell type proportions *π*_**1**_, *π*_**2**_, then average to get 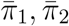.

For estimation of the parameters {*ω*_1_, *ω*_2_, *ω*_12_, *c*_1_, *c*_2_, *c*_3_, *c*_12_, *c*_13_, *c*_23_}, based on Eq. (S7), consider the regressions

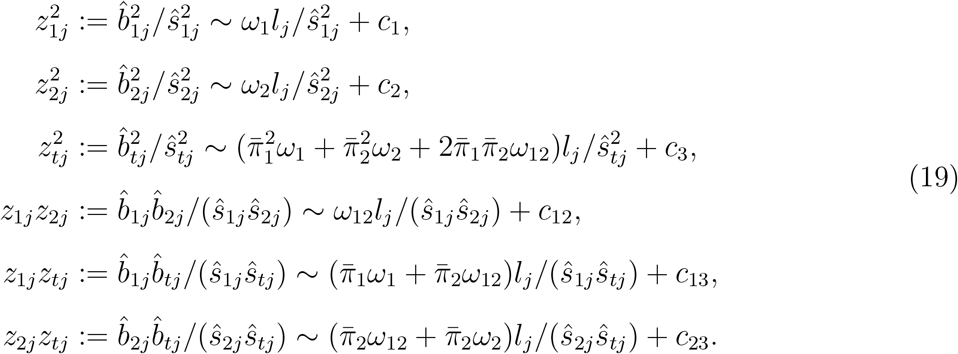

We separately regress 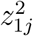 on 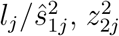 on 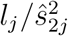 and *z*_1*j*_*z*_2*j*_ on 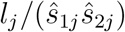. The slopes 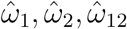 and intercepts *ĉ*_1_, *ĉ*_2_, *ĉ*_12_ are estimates of *ω*_1_, *ω*_2_, *ω*_12_ and *c*_1_, *c*_2_, *c*_12_, respectively. Also, the inter-cepts *ĉ*_3_, *ĉ*_13_, *ĉ*_23_ of regressing 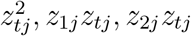 on 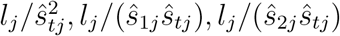 give the estimates of *c*_3_, *c*_13_, *c*_23_, respectively. In practice, if there are no sample overlap, the matrix **C** can be set as a diagonal matrix to alleviate the burden of parameter estimation, making the estimation of (co-) heritability more stable. Besides, since the cis-SNPs of a gene are typically in a small number (tens to hundreds), to avoid unreasonable co-heritability estimates, we adopt a conservative approach: setting 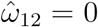 if *p*-value of the Wald test 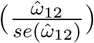 is smaller than a threshold (e.g 0.05).

Finally, we directly plug the estimated parameters into Eq. (17) and Eq. (18) to obtain IBSEP estimators 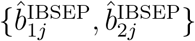 and their variances 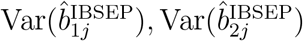. Cell-type-specific cis-SNPs with non-zero effects on the target gene can be detected using Wald test based on IBSEP 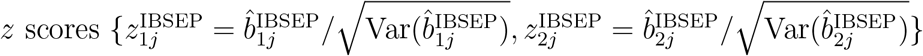.

### Experimental validation on the usage of mean cell type proportions

Following the simulation setting in Eq. (2), the true gene expression vector at the tissue level can be generated using the formula **y**_*t*_ = ***π***_1_ ⊙ **X**_*t*_***β***_1_ + ***π***_2_ ⊙ **X**_*t*_***β***_2_ + ***ϵ***_*t*_, where ***π***_1_ and ***π***_2_ are the individual proportions of the two cell types, we thereforely denoted this **y**_*t*_ as 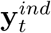. By contrast, the approximation theory from Eq. (8) allows us to generate an approximate expression 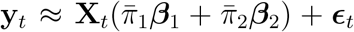, we denoted this **y**_*t*_ as 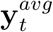 because of the usage of average cell type proportions.

After obtaining the simulated gene expression vectors 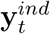 and 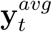, they are independently associated with the genotype data **X**_*t*_ to obtain marginal effect size estimates 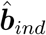 and 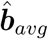, respectively, which are then compared. As shown in Fig. S1, even with high heterogeneity in ***π***_1_ and ***π***_2_ (i.e., *α*=6), the average Pearson correlation coefficient between **b**_*ind*_ and **b**_*avg*_ significantly increases with the sample size *N*_*t*_ increase, consistent with the law of large numbers. Moreover, as expected, when the heterogeneity in ***π***_1_ and ***π***_2_ are reduced (i.e., *α*=50), the Pearson correlation coefficient further increases, reaching 0.99978 (s.e.=0.00001) when *N*_*t*_=100. Fixing *N*_*t*_=1000, it can be observed that the Pearson correlation coefficient significantly improves as the heterogeneity in ***π***_1_ and ***π***_2_ decreases.

### Compared methods in simulation

In the simulation of type I error and power evaluation, the compared methods include “Cell type 1”, “Cell type 1&2” and “Tissue ieQTL”. “Cell type 1” only uses the summary statistics of cell type 1 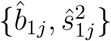. “Cell type 1&2” uses the summary statistics of cell type 1 and cell type 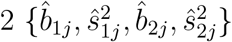 which can be viewed as a special case of IBSEP without tissue eQTLs. The estimator of “Cell type 1&2” is derived in a similar as the full version of IBSEP. Without tissue-level eQTL summary statistics, the conditional mean of the estimated marginal effects and conditional variance become:

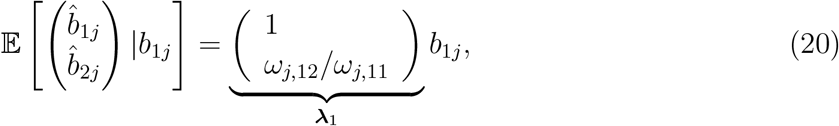

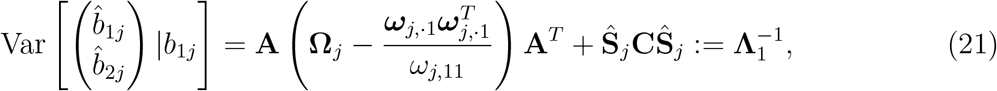

where

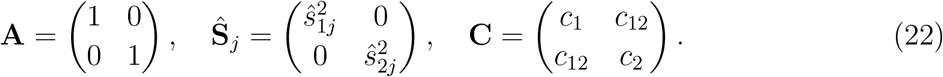

The IBSEP-ct estimator and corresponding variance are as follows:

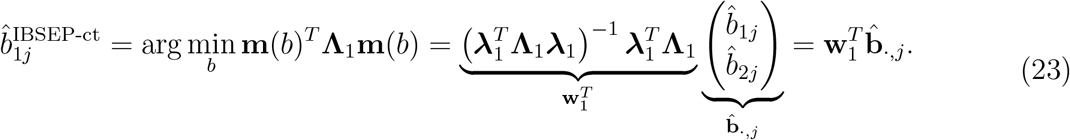

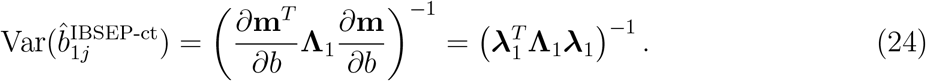

For “Tissue ieQTL”, we follow the commonly used approach to test the interaction between tissue-level gene expression and cell type proportion [9]. The linear mixed model with an interaction term is:

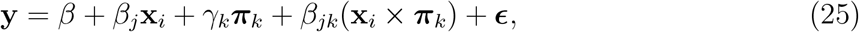

where **x**_*j*_ ∈ *ℝ*^*N*^ is the genotype vector of SNP *j*, ***π***_*k*_ ∈ *ℝ*^*N*^ is the proportion vector of cell type *k* and (**x**_*i*_ *×* ***π***_*k*_) is the interaction term. We test *β*_*jk*_ = 0: *β*_*jk*_ *≠* 0 means SNP *j* is an ieQTL of cell type *k*.

### Data preparation in real data analysis

For blood ct-eQTL analysis, variants in 1000 Genomes project reference panel with minor allele frequency (MAF) *<* 1% were excluded. We merged summary statistics of GTEx whole blood, each cell type of 1M and each sub-cell type of OneK1K with the reference panel by SNPs passed the quality control and aligned the effect allele. We retained genes that were shared in all the six cell types of 1M and the 14 sub-cell types of OneK1K. For each SNP, we calculated its LD score with SNPs mapped to the same gene using LDSC package. Since 1M only considered SNPs within a 100kb distance to each gene midpoint, we excluded genes with fewer than 50 SNPs, resulting in 5302 genes.

For brain ct-eQTL analysis, a similar data processing procedure was adopted. We merged summary statistics of GTEx brain cortex and each cell type of sc-brain with 1000 Genomes project reference panel by SNPs. Considering the significant differences in the genes contained within different cell types of sc-brain (12831 genes in excitatory neurons, 5871 genes in pericytes), we did not take the intersection of genes across all the eight cell types. As sc-brain mapped cis-eQTLs within a 1-Mb window of the TSS of a gene, each gene typically contains thousands of SNPs. We thereforely did not further filter genes.

### Colocalization

The colocalization analysis was conducted following the procedure described in [13]. For AD, BIP, MDD and SCZ, we defined the coordinates of SNPs in LD (*r*^2^ *>* 0.1 in 1000 Genome European cohort) with the reported lead SNPs as loci using LDlinkR [87]. For RA, MS, IBD and asthma, we defined lead SNP as the SNP passing the genome-wide significant threshold (*p*-value *<* 5 *×* 10^−8^) with the most significant *p*-value in each cytoband. For each loci, we tested colocalization between the GWAS and eQTL signals for genes with at least 10 SNPs overlapping with this loci using the “coloc.abf” function of the *Coloc* R package [31]. “coloc.abf” outputs “PP.H4.abf” which is the posterior probability that the gene and the disease share a same causal SNP. We defined colocalized genes as those with “PP.H4.abf” greater than 0.7 and colocalized loci as those containing at least one colocalized gene.

## Data Availability

All data produced in the present work are contained in the manuscript

## Data Availability

Blood cell-type level eQTL summary statistics are available at https://molgenis26.gcc.rug.nl/downloads/1m-scbloodnl/eqtls_20201106_genome_wide.tar.gz (1M) and https://onek1k.org/ (OneK1K). Brain cell-type level eQTL summary statistics are available at https://zenodo.org/records/7276971 (sc-brain). GTEx tissue-level eQTL summary statistics are available at https://storage.googleapis.com/gtex-resources/GTEx_Analysis_v8_QTLs/GTEx_Analysis_v8_eQTL_all_associations/Whole_Blood.allpairs.txt.gz (whole blood) and https://storage.googleapis.com/gtex-resources/GTEx_Analysis_v8_QTLs/GTEx_Analysis_v8_eQTL_all_associations/Brain_Cortex.allpairs.txt.gz (brain cortex). GTEx tissue expressions are available at: https://storage.googleapis.com/adult-gtex/bulk-gex/v8/rna-seq/counts-by-tissue/gene_reads_2017-06-05_v8_whole_blood.gct.gz (whole blood) and https://storage.googleapis.com/adult-gtex/bulk-gex/v8/rna-seq/counts-by-tissue/gene_reads_2017-06-05_v8_brain_cortex.gct.gz (brain cortex). 1000 Genomes reference panels can be downloaded at https://console.cloud.google.com/storage/browser/broad-alkesgroup-public-requester-pays;tab=objects?prefix=&forceOnObjectsSortingFiltering=false. GWAS summary statistics are available here: Rheumatoid arthritis: https://data.cyverse.org/dav-anon/iplant/home/kazuyoshiishigaki/ra_gwas/ra_gwas-10-28-2021.tar; Multiple sclerosis: https://imsgc.net/; Inflammatory bowel disease: ftp://ftp.sanger.ac.uk/pub/project/humgen/summary_statistics/human/2016-11-07/; Asthma: https://broad-ukb-sumstats-us-east-1.s3.amazonaws.com/round2/additive-tsvs/20002_1111. gwas.imputed_v3.both_sexes.tsv.bgz; Alzheimer’s disease: http://ftp.ebi.ac.uk/pub/databases/gwas/summary_statistics/GCST90012001-GCST90013000/GCST90012877/GCST90012877_buildGRCh37.tsv.gz; Bipolar disease: https://figshare.com/ndownloader/files/26603681; Major depressive disease: https://datashare.ed.ac.uk/bitstream/handle/10283/3203/PGC_UKB_depression_genome-wide.txt?sequence=3&isAllowed=y; Schizophrenia: https://figshare.com/ndownloader/files/34517828.

## Code Availability

The IBSEP software and scripts for reproducing the analyses are available at: https://github.com/xinyiyu/IBSEP.

