## Supplementary information for "A unified framework for cell-type-specific eQTLs prioritization by integrating bulk and scRNA-seq data"

### 1 Supplementary Figures

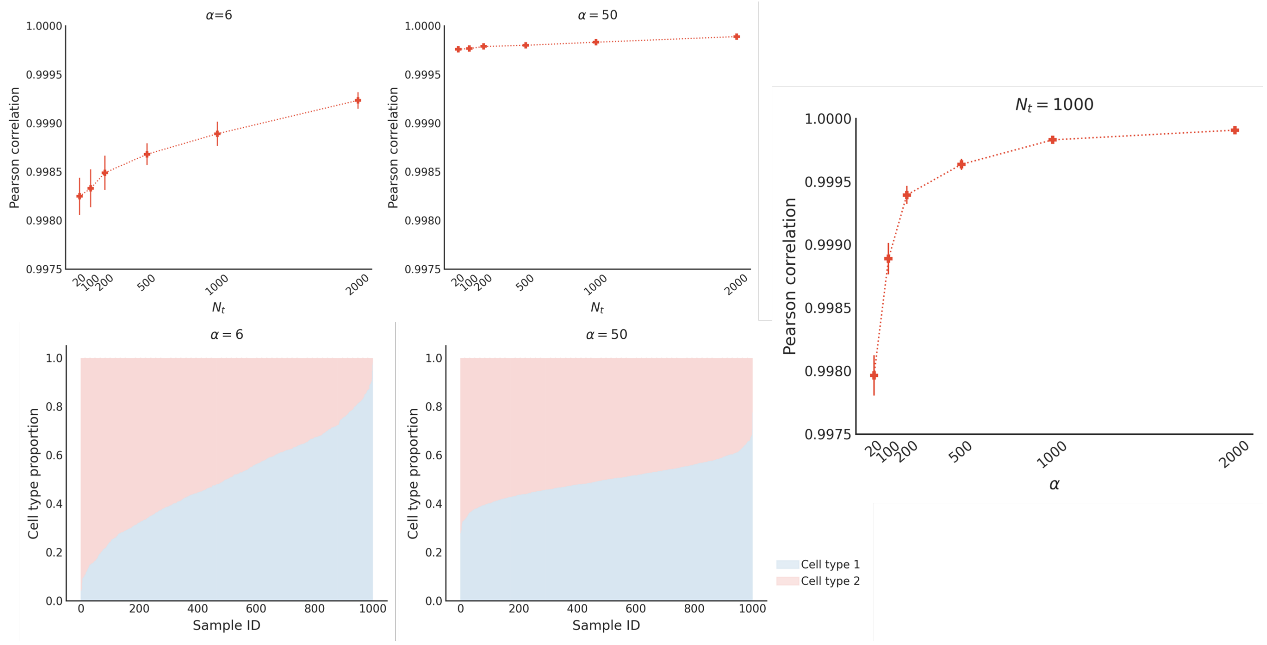

Figure S1: **Validation of using mean cell type proportions**. We randomly chose $N_t$ individuals from UKBB and used the genotype of the first LD window in chromosome 20. The individual cell type 1 proportions $\pi_{1j}$'s were from Beta$(\alpha\bar{\pi}_1, \alpha(1-\bar{\pi}_1))$, where a smaller $\alpha$ leads to larger variation. $\boldsymbol{\beta}_1, \boldsymbol{\beta}_2$ followed Eq. (1) in the main paper. We set $\bar{\pi}_1 = 0.5$, $\alpha \in \{6, 50\}$, $p_{cau} = 0.05$, $h_1^2 = h_2^2 = 0.05$. The gene expression $\mathbf{y}_t$ was generated by (i) Eq. (2) and (ii) the tissue model in Eq. (3), respectively. We then calculated the marginal effect sizes $\boldsymbol{\beta}_t$ of (i) and (ii). The stacked barplots show the individual cell type proportions with different $\alpha$, and the upper line plots show the corresponding average Pearson correlations between the marginal effect sizes of (i) and (ii) with various tissue sample sizes ($N_t$). The line plot on the right show the change of average Pearson correlations with varying $\alpha$. The error bars are standard deviations of 50 replications.

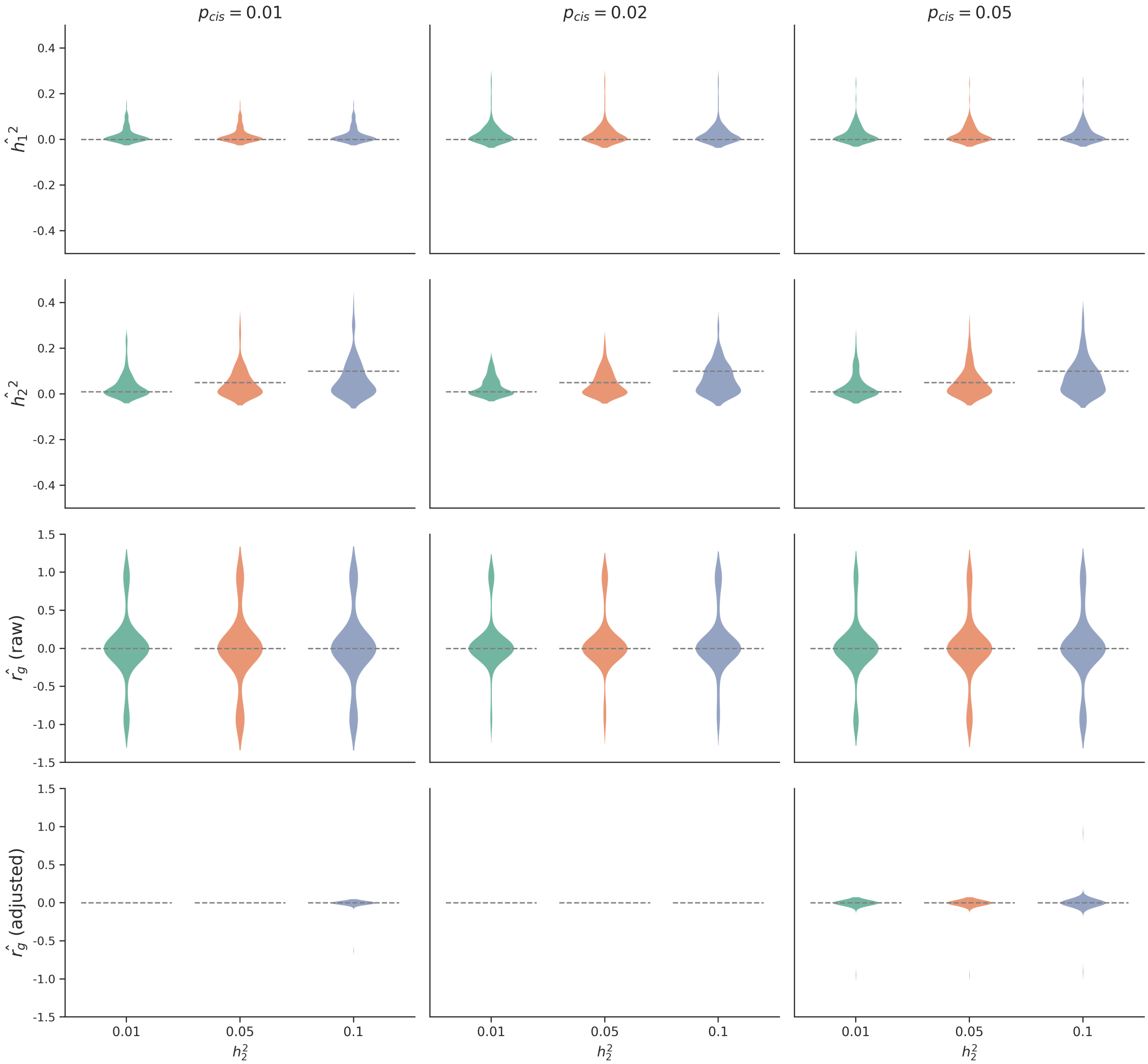

Figure S2: **Parameter estimation in the type I error evaluation simulation.**

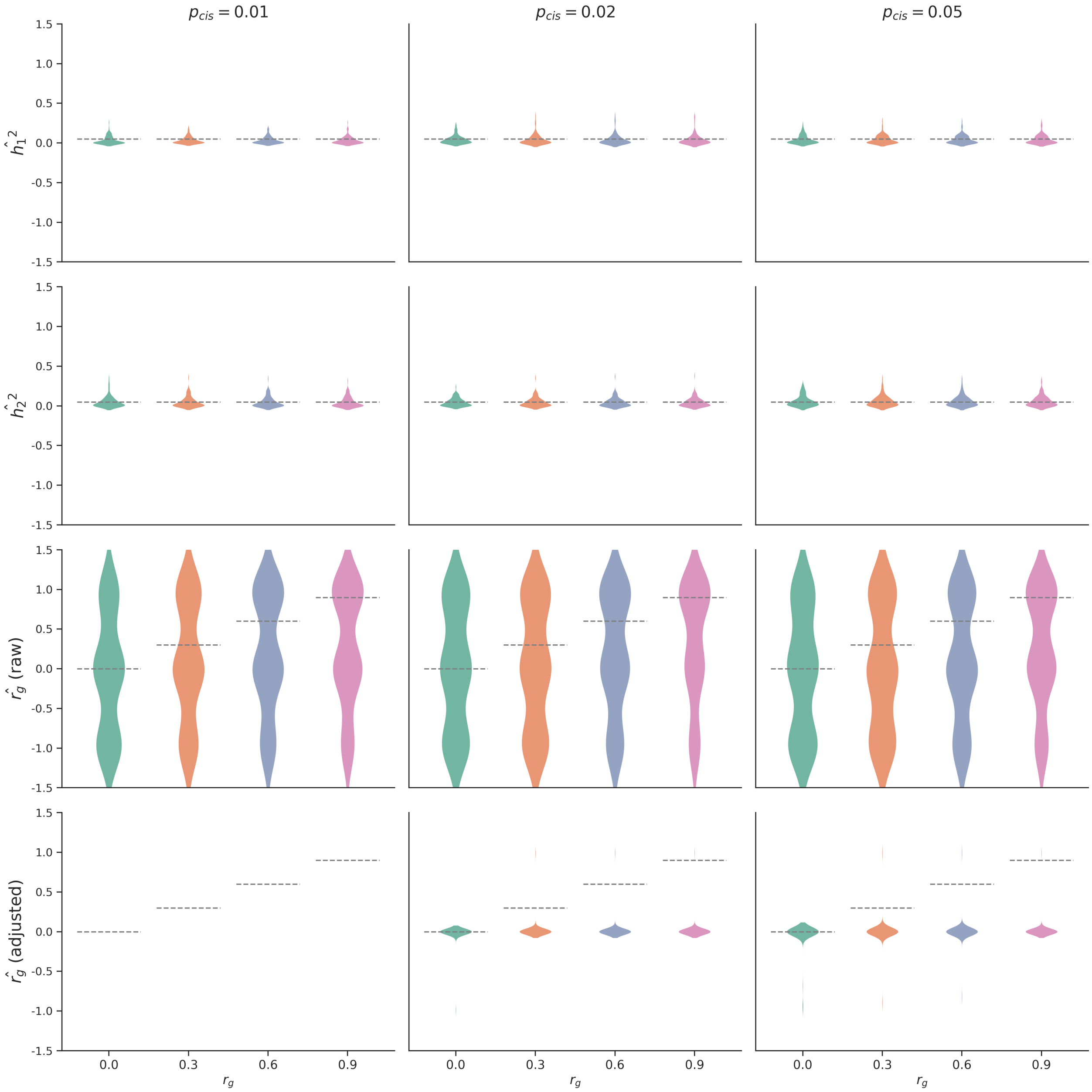

Figure S3: **Parameter estimation in the power evaluation simulation.**

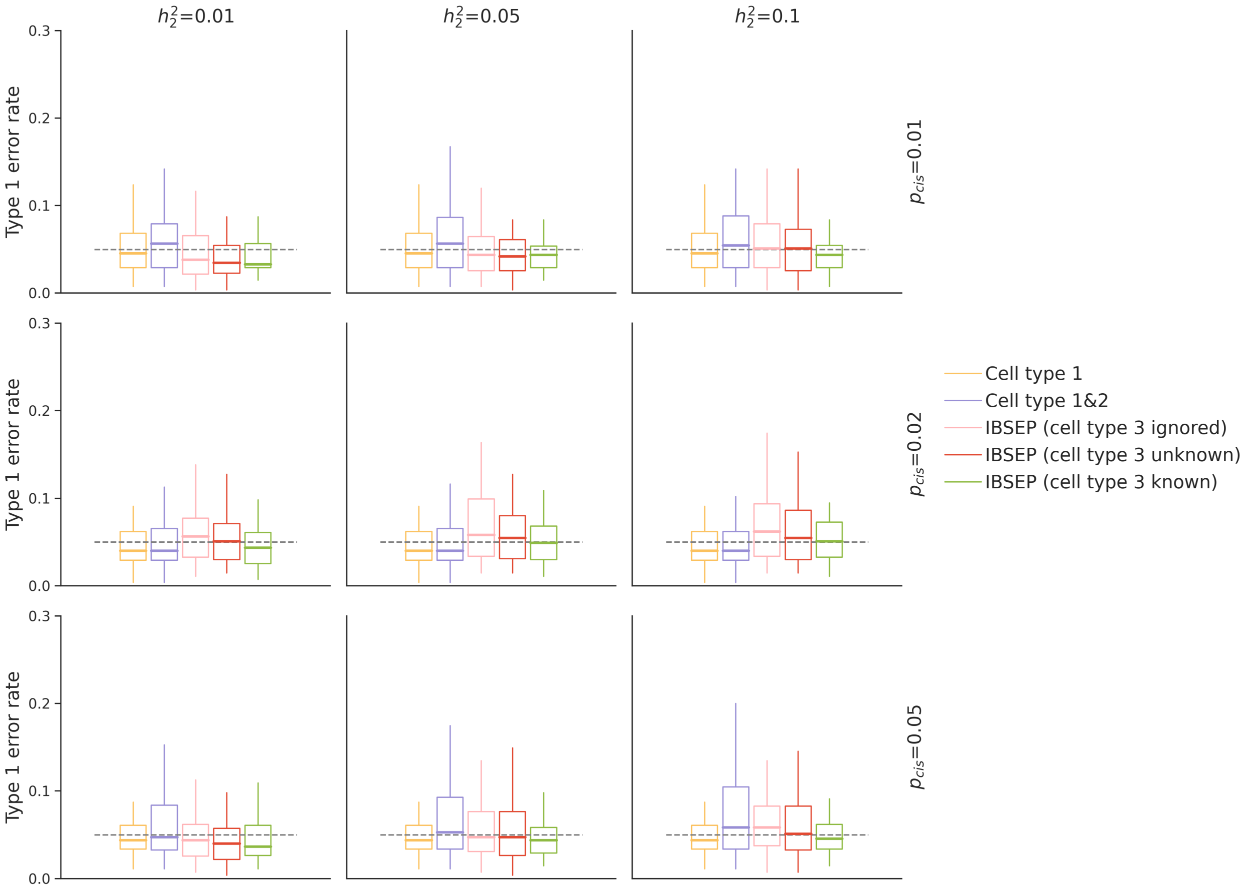

Figure S4: **Type I error rate assessment when a latent cell type is present**. Data of cell type 1, 2 and 3 was generated in a same way as in the main type I error simulation. The tissue gene expression is generated by $\mathbf{y}_t = \boldsymbol{\pi}_1 \odot \mathbf{X}_t\boldsymbol{\beta}_1 + \boldsymbol{\pi}_2 \odot \mathbf{X}_t\boldsymbol{\beta}_2 + \boldsymbol{\pi}_3 \odot \mathbf{X}_t\boldsymbol{\beta}_3 + \boldsymbol{\epsilon}_t$. We set the true mean cell type proportions $\pi_1 = 0.4, \pi_2 = 0.4, \pi_3 = 0.2$. IBSEP was performed in three ways: (i) ignoring the existence of cell type 3 in the tissue data, i.e., used unnormalized cell type proportions $\pi_1, \pi_2$, then applied the mis-specified model with 2 cell types; (ii) being unaware of the existence of cell type 3 in the tissue data, i.e., used the normalized cell type proportions $\frac{\pi_1}{\pi_1+\pi_2}, \frac{\pi_2}{\pi_1+\pi_2}$, then applied the mis-specified model with 2 cell types; (iii) applying the correct model with 3 cell types.

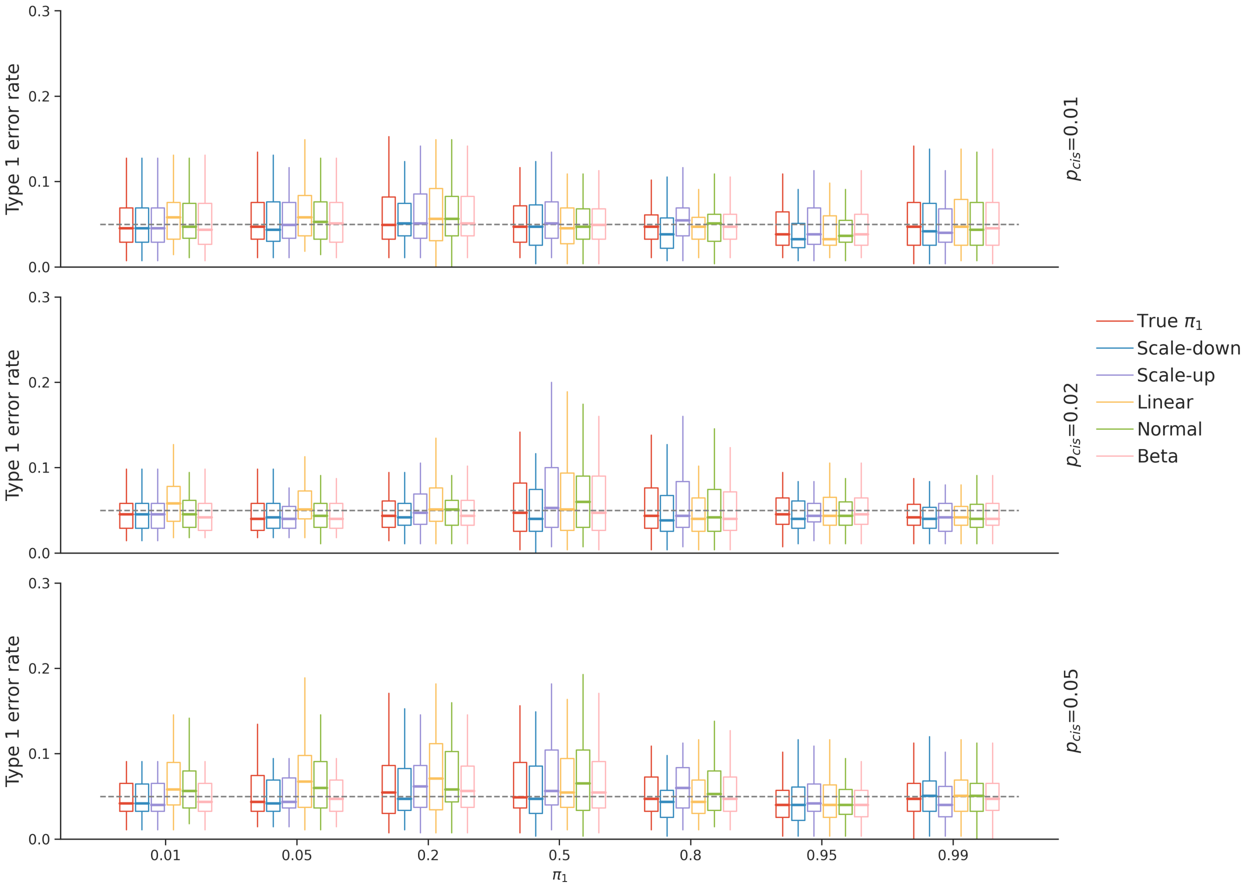

Figure S5: **Type I error rate assessment when the mean cell type estimation is inaccurate**. Different from the main type I error simulation where the true mean cell type proportions were plugged in the IBSEP estimator, here we used several types of inaccurate $\hat{\pi}_1$ (thus inaccurate $\hat{\pi}_2$): (i) $\hat{\pi}_1 = 0.8\pi_1$ ("scale-down"); (ii) $\hat{\pi}_1 = 1.2\pi_1$ ("scale-up"); (iii) $\hat{\pi}_1 = 0.7\pi_1 + 0.2$ ("linear"); (iv) $\hat{\pi}_1 \sim \mathcal{N}(\pi_1, 0.05^2)$ ("normal"); (v) $\hat{\pi}_1 \sim \text{Beta}(50\pi_1, 50(1-\pi_1)$ ("beta").

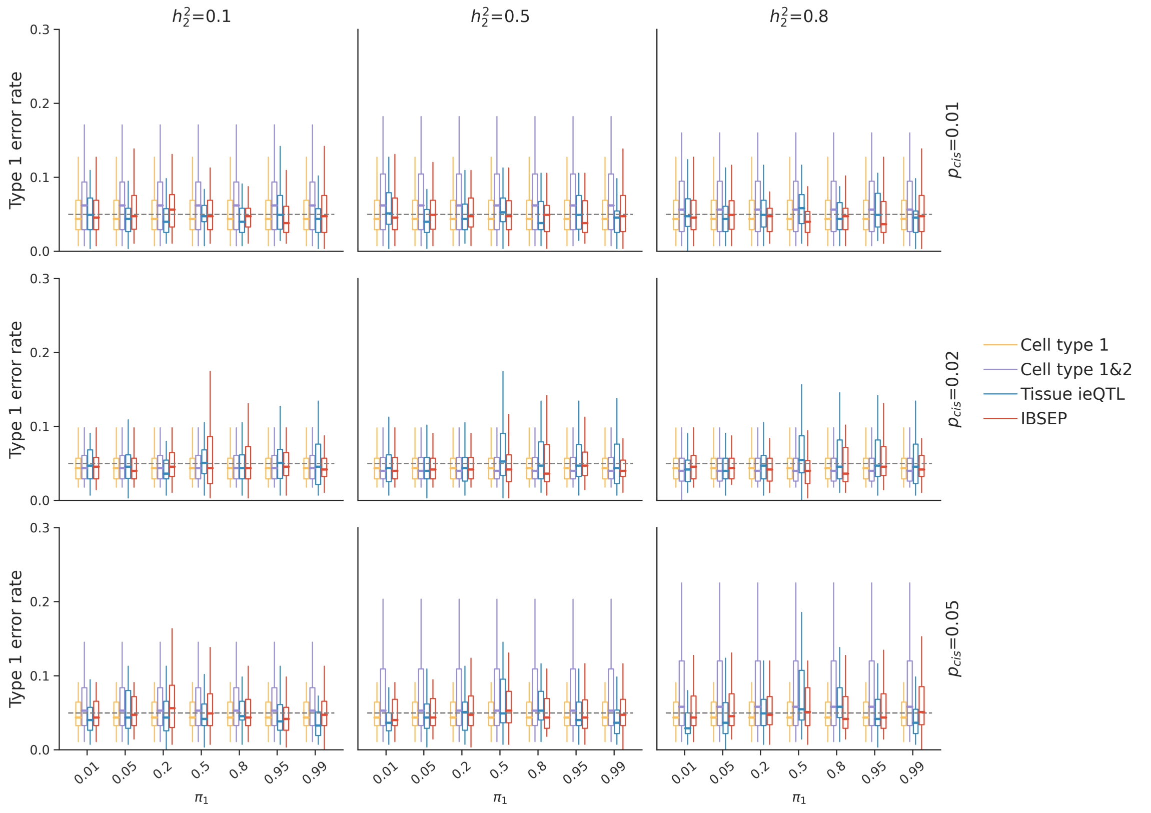

(a) Type I error rate assessment with higher heritability.

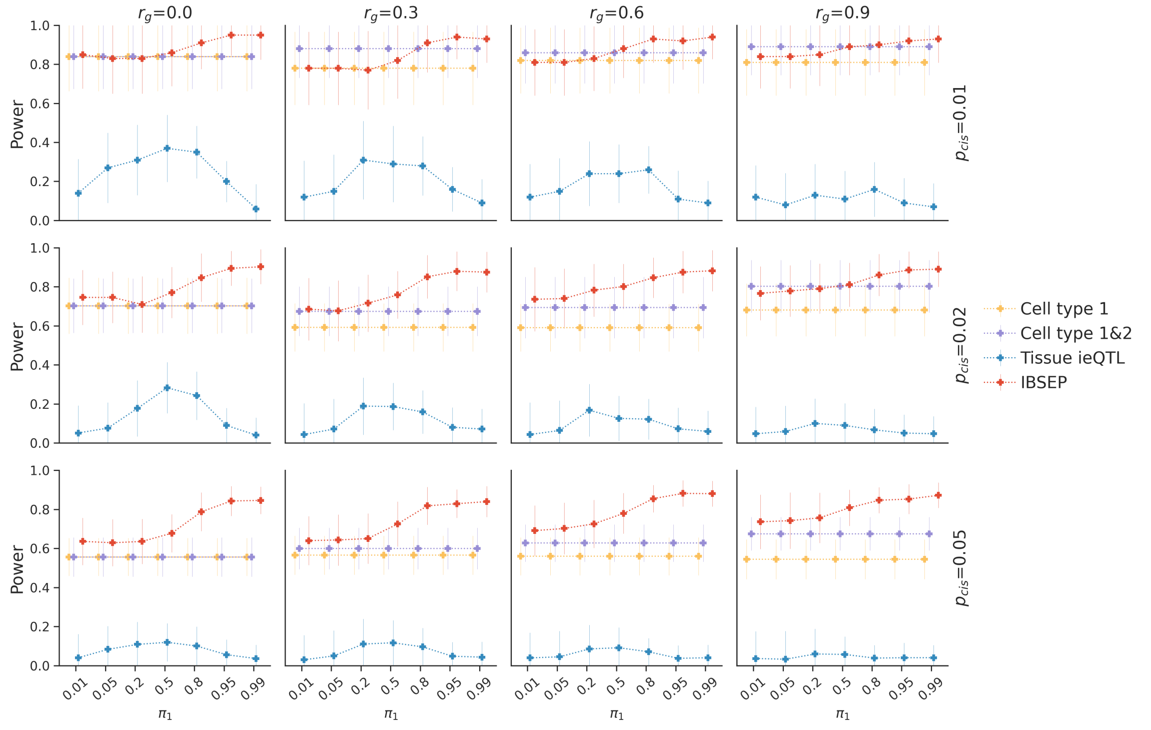

(b) Power evaluation with $h_2 = 0.8$.

Figure S6: **Simulation with higher heritability to investigate the performance of ieQTL.**

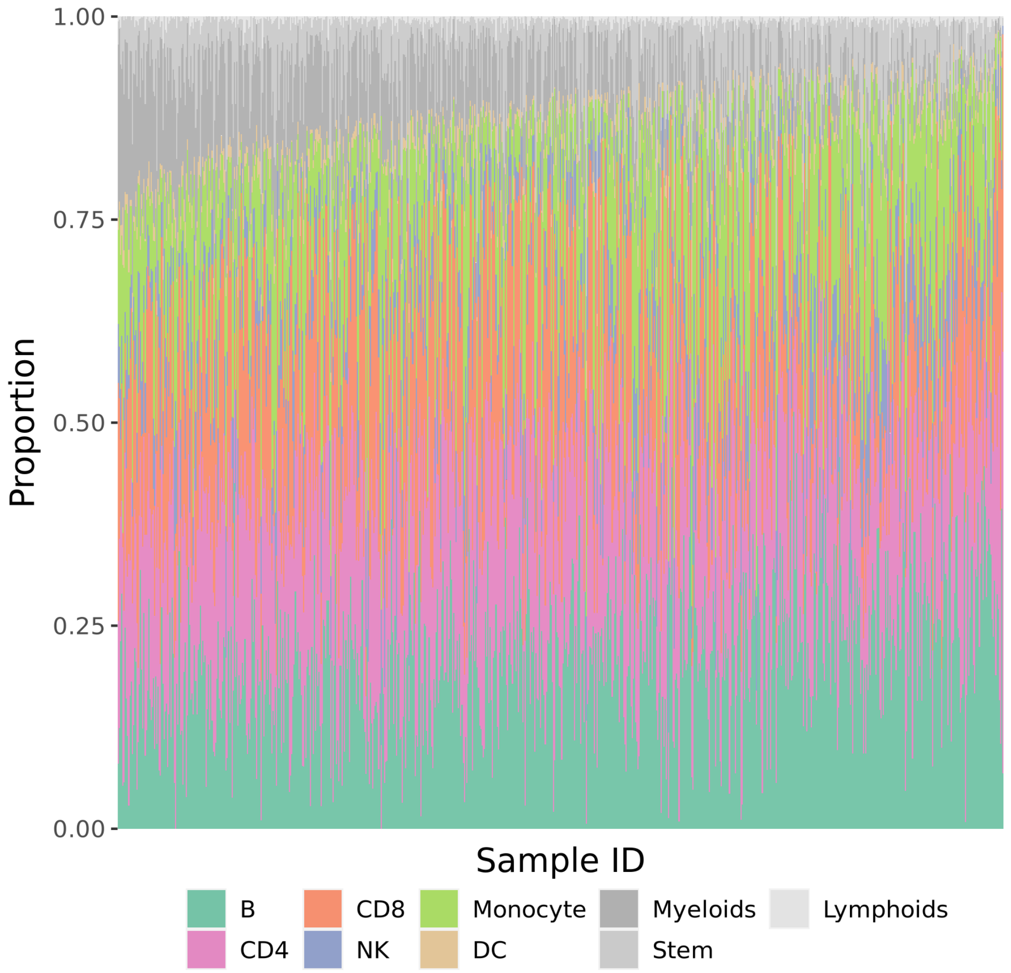

Figure S7: **Cell type compositions of GTEx blood samples**. xCell estimates proportions of 64 cell types including the six studied cell types (B, CD4, CD8, monocyte, NK and DC) and other cell types. Cell types provided by xCell that are not among the six studied cell types are categorized into myeloid, stem, and lymphoid cells.

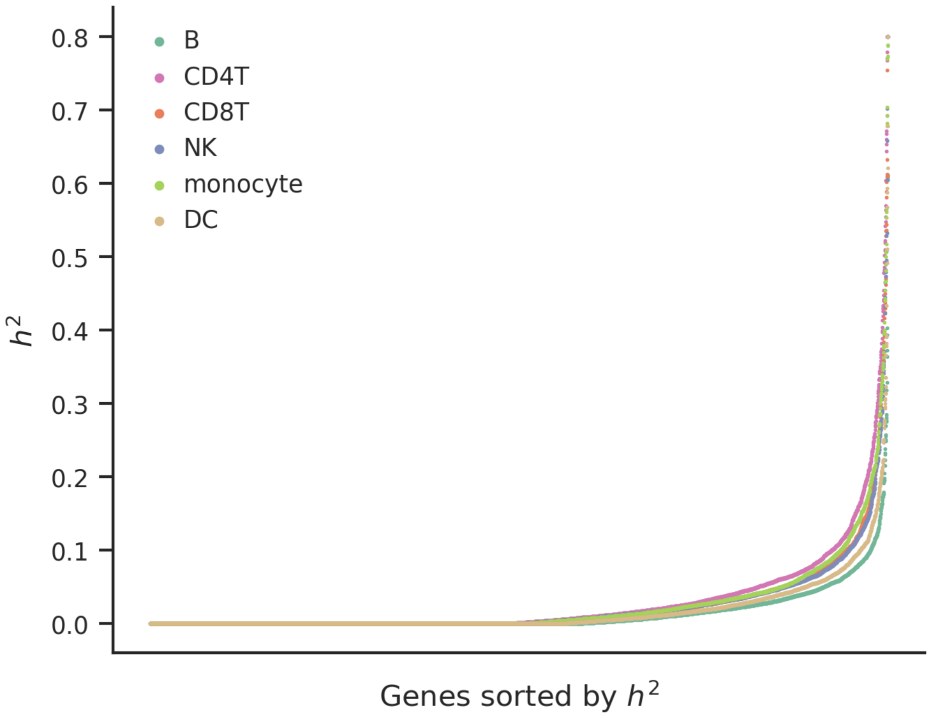

Figure S8: **Cis-heritability estimation in each blood cell type.** For each gene and cell type $i$, $i = 1, ..., 6$, cis-heritability estimate $\hat{\omega}_{ii}$ is obtained from LD score regression.

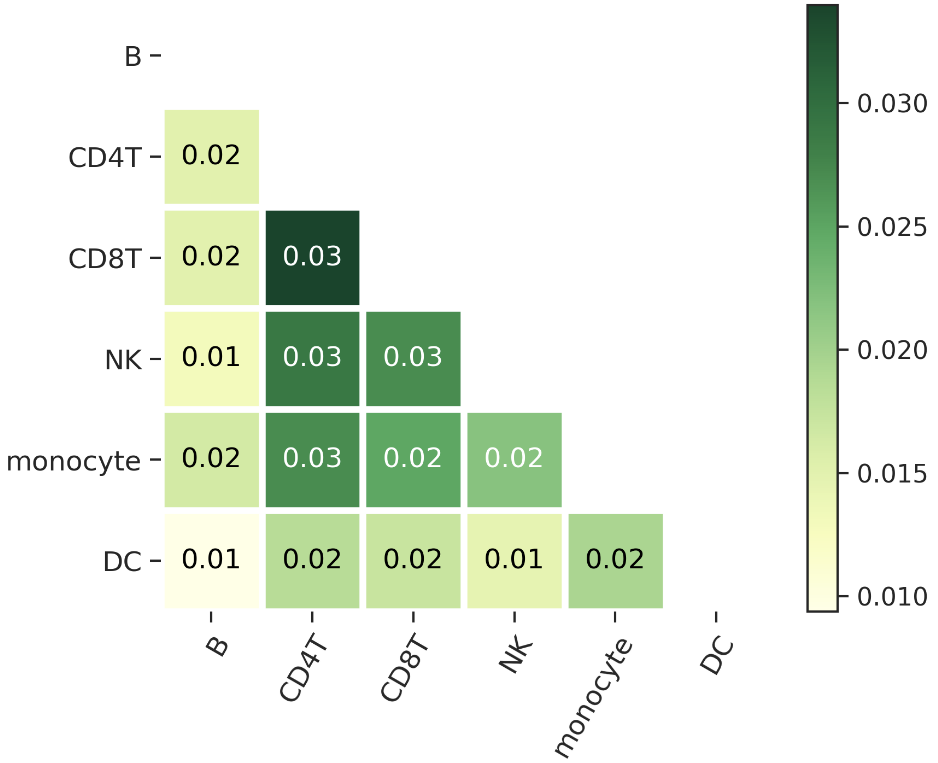

Figure S9: **Heatmap shown the fraction of genes with significant genetic correlation among cell types.** For each pair of cell types, we calculate the fraction of genes with significant genetic correlation.

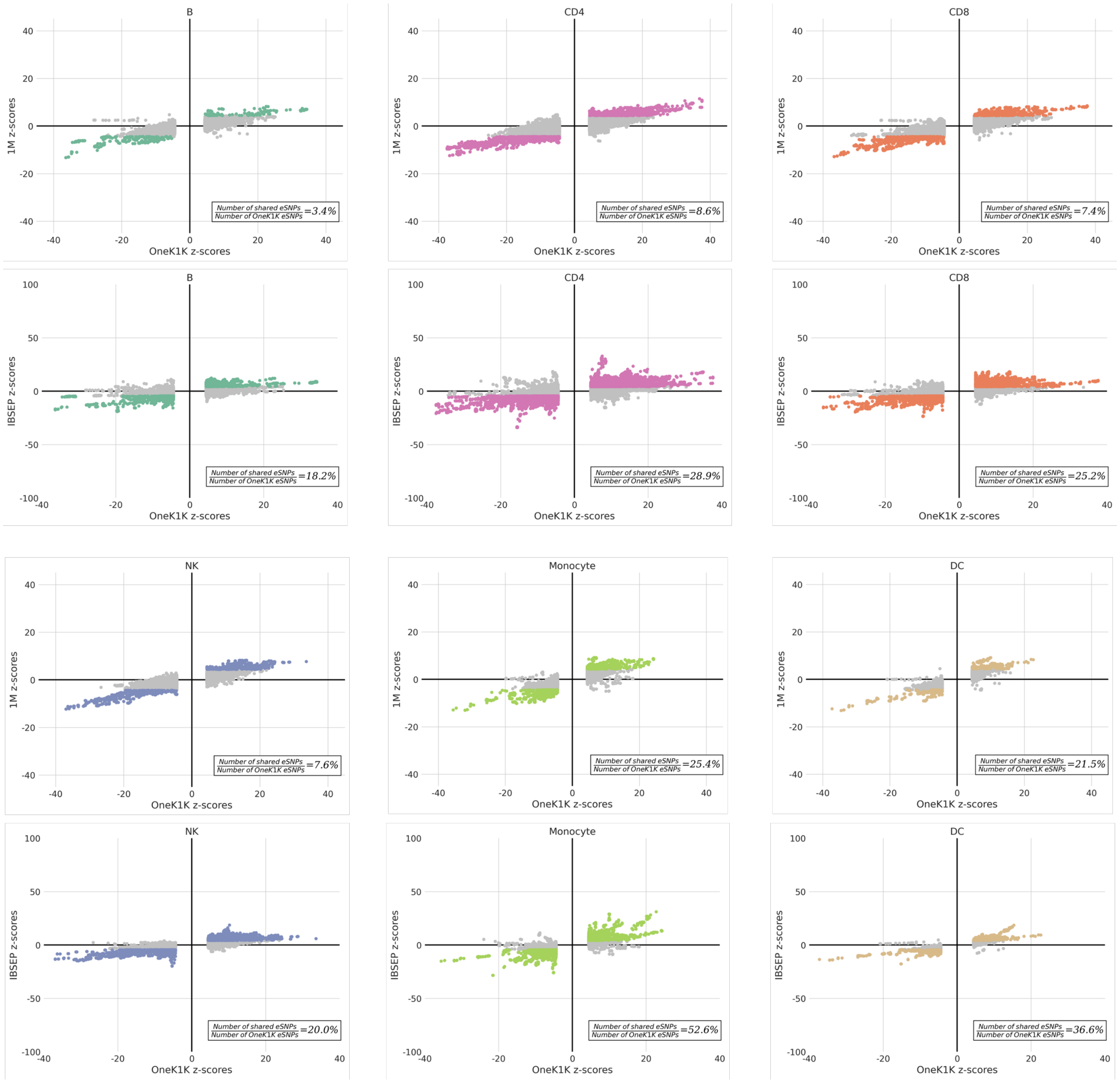

Figure S10: **Comparison of $z$-scores of OneK1K eSNPs with 1M and IBSEP.** For each cell type, the upper plot shows the comparison of $z$-scores between OneK1K eSNPs and the same SNPs in 1M; the lower plot shows the comparison of $z$-scores between OneK1K eSNPs and the same SNPs in IBSEP.

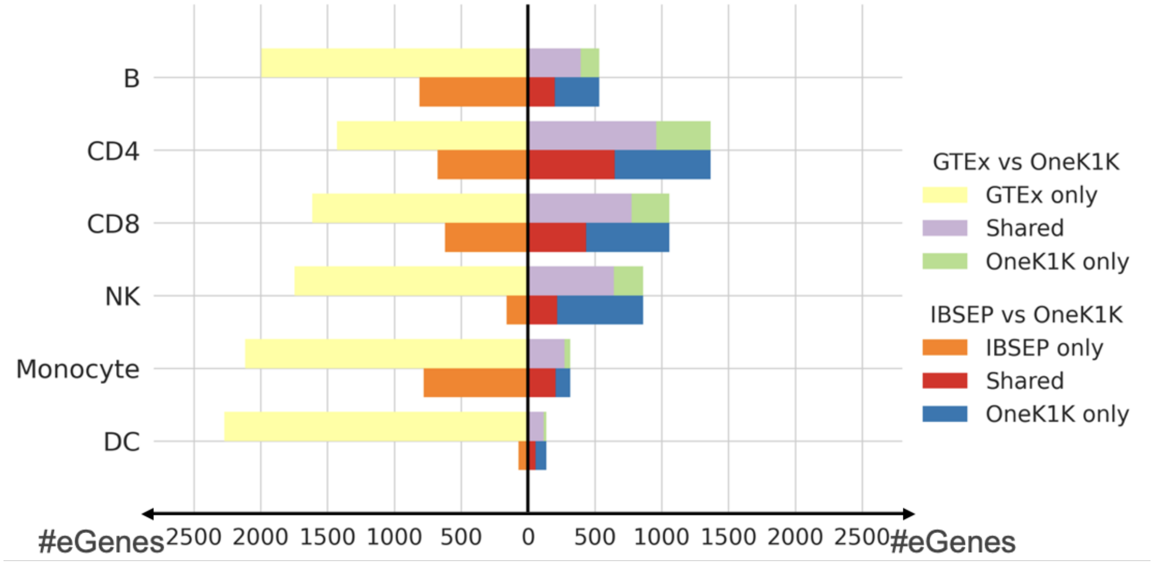

Figure S11: **Barplot comparing the number of eGenes discovered by GTEx and IBSEP respectively with OneK1K in each cell type.**

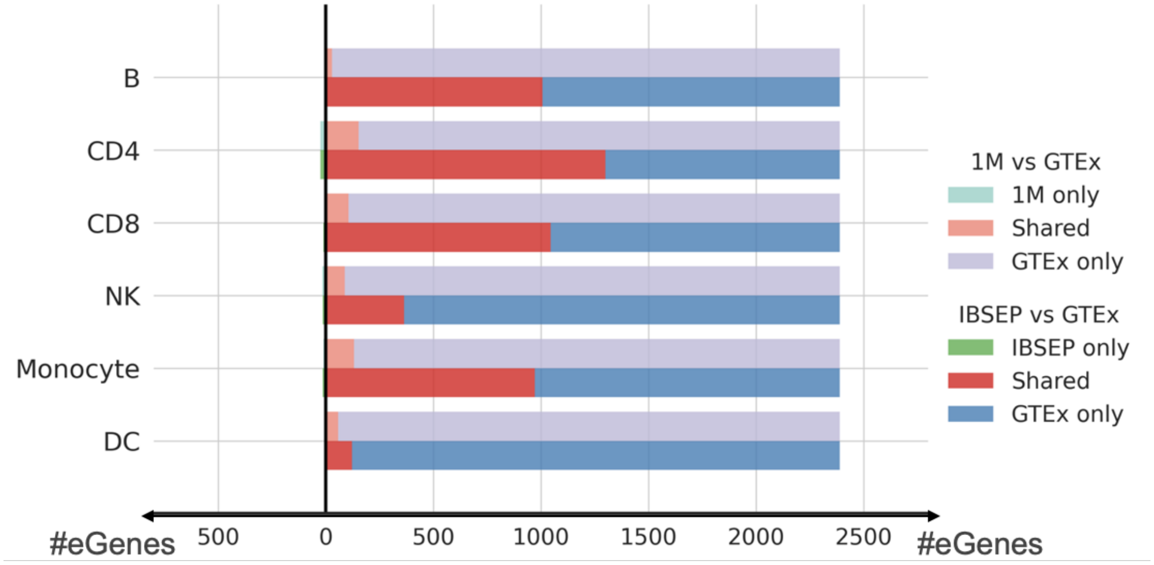

Figure S12: **Barplot comparing the number of eGenes discovered by 1M and IBSEP respectively with GTEx in each blood cell type.**

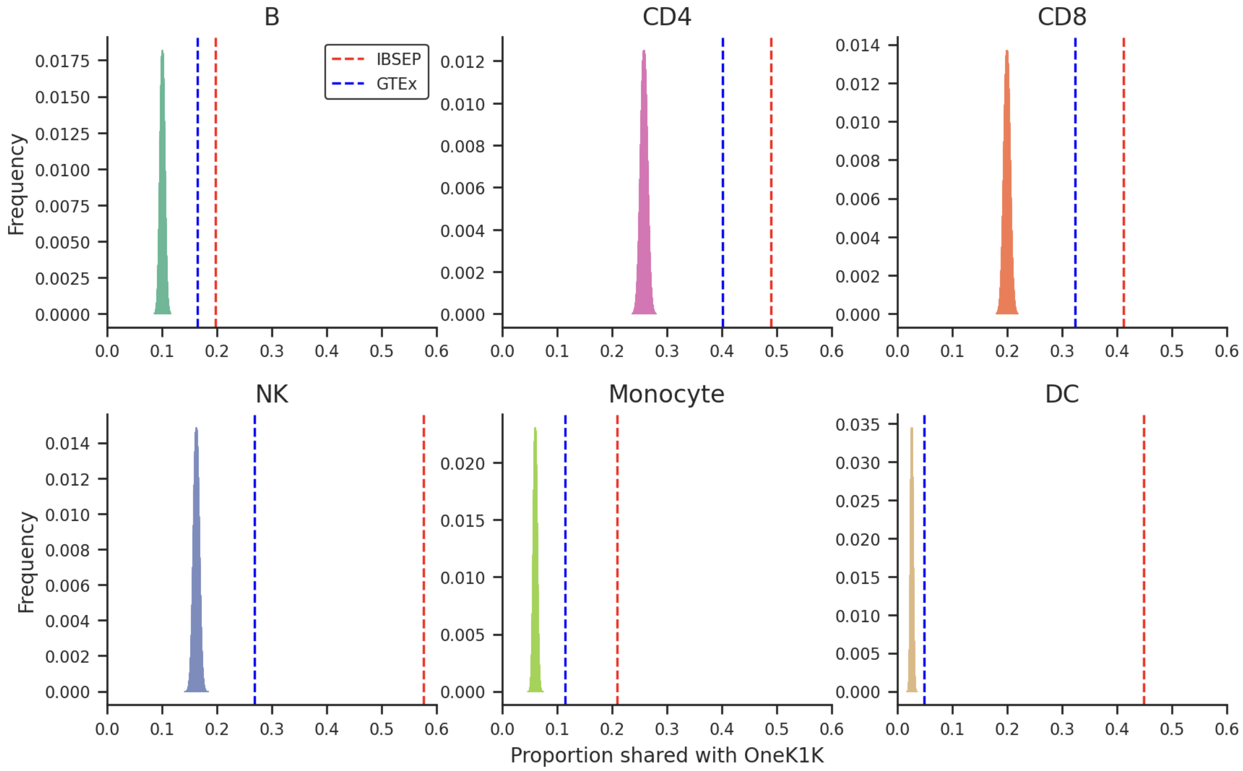

Figure S13: **Comparison of the fraction of eGenes shared with OneK1K between IBSEP and GTEx in each cell type.** For each cell type, the red and blue dashed line respectively represent the proportion of eGenes shared with OneK1K among all eGenes discovered by IBSEP and GTEx. The histogram represents the distribution of the proportion of overlap with OneK1K eGenes if randomly selecting the same number of genes from all genes as IBSEP eGenes.

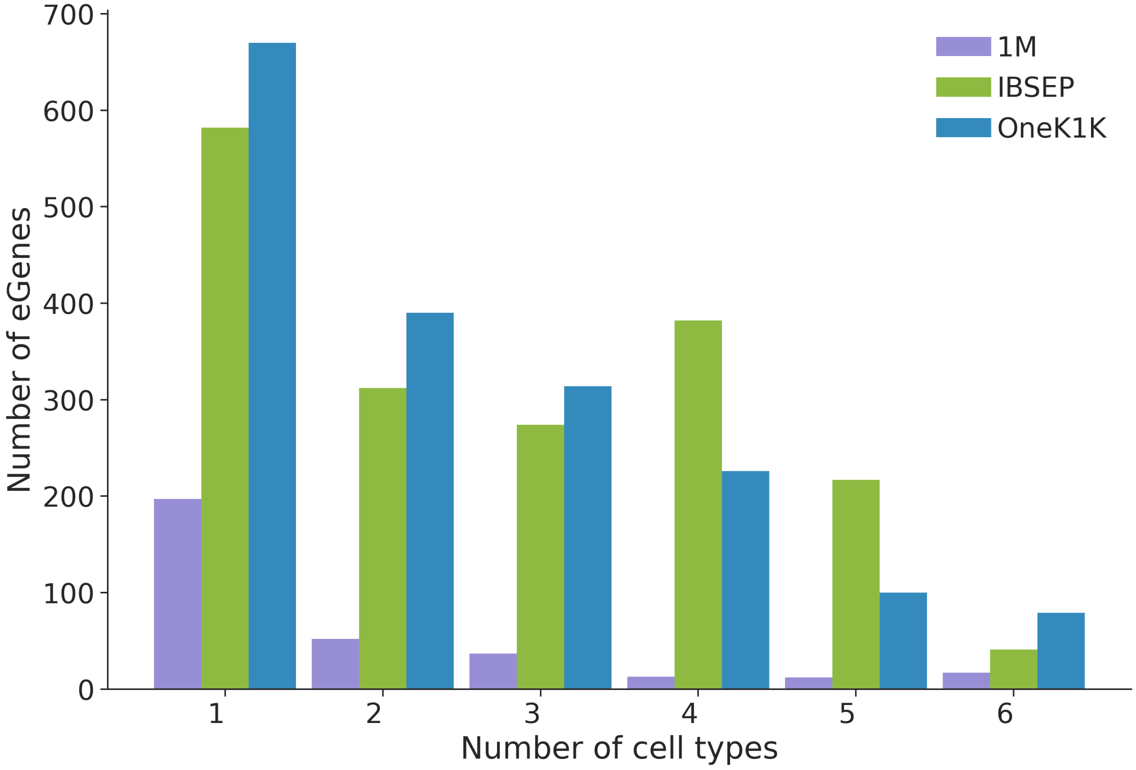

Figure S14: **Number of cell types in which the eGene is identified by 1M, IBSEP and OneK1K.**

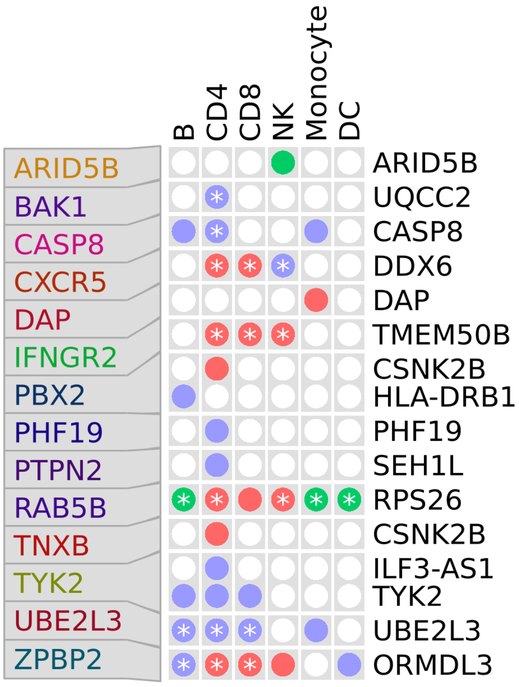

Figure S15: **Cell-type-level colocalized genes of rheumatoid arthritis found by sc-brain and IBSEP.**

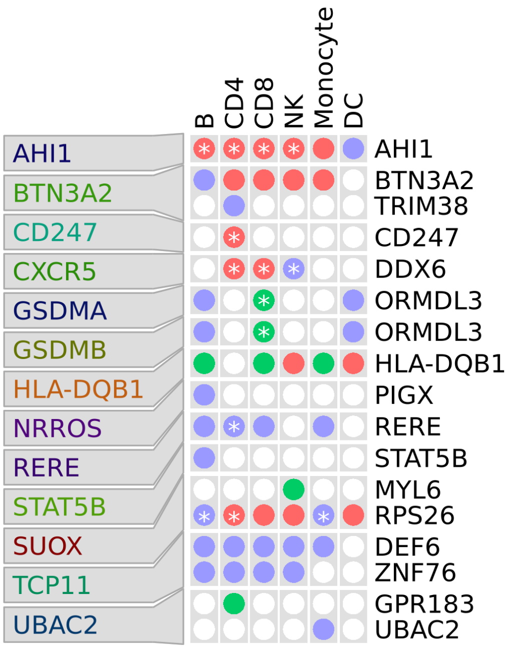

Figure S16: **Cell-type-level colocalized genes of asthma found by sc-brain and IBSEP.**

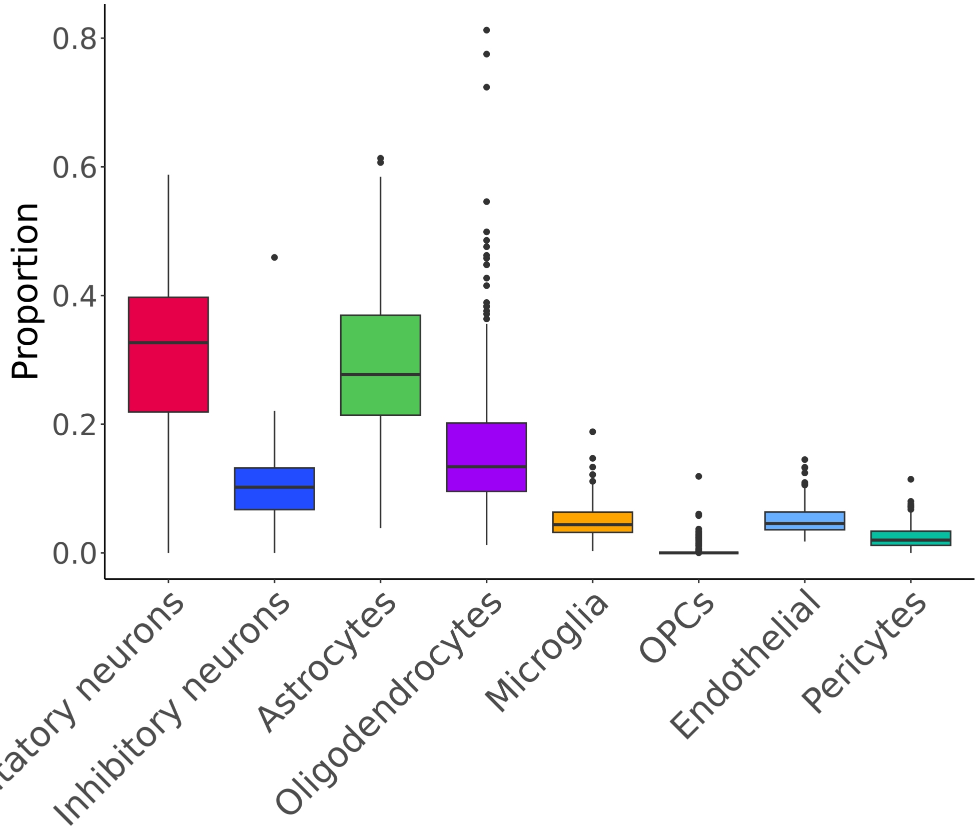

Figure S17: **Cell type proportions of GTEx brain cortex samples**.

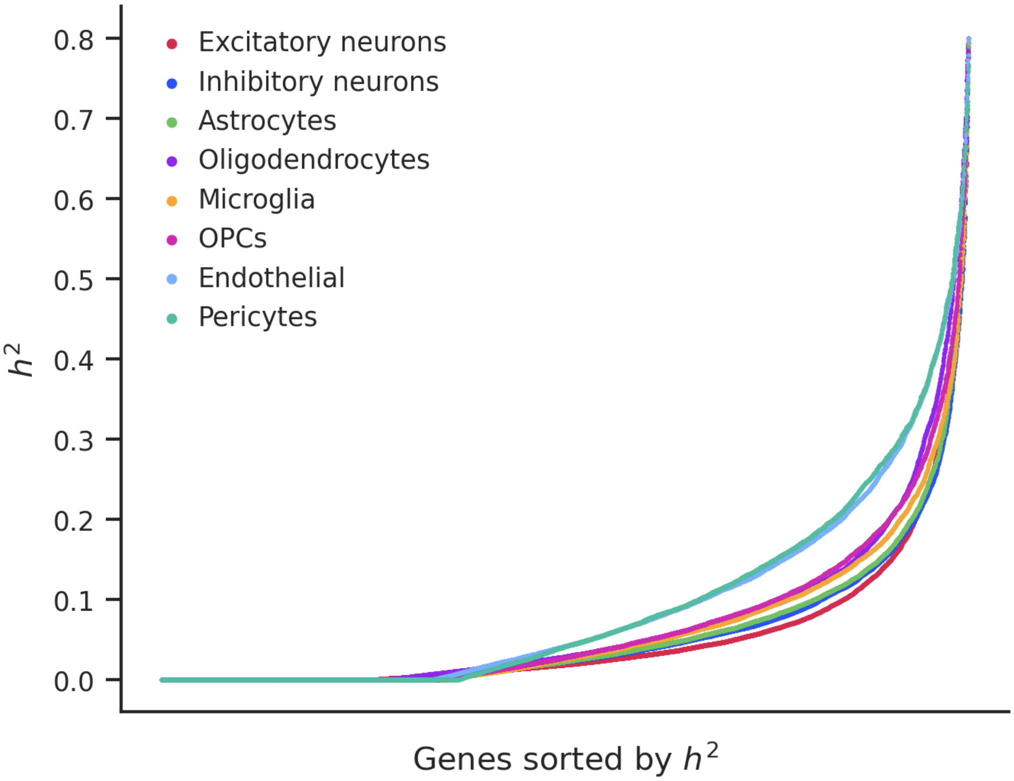

Figure S18: **Cis-heritability estimation in each brain cell type.** For each gene and cell type $i$, $i = 1, ..., 6$, cis-heritability estimate $\hat{\omega}_{ii}$ is obtained from LD score regression.

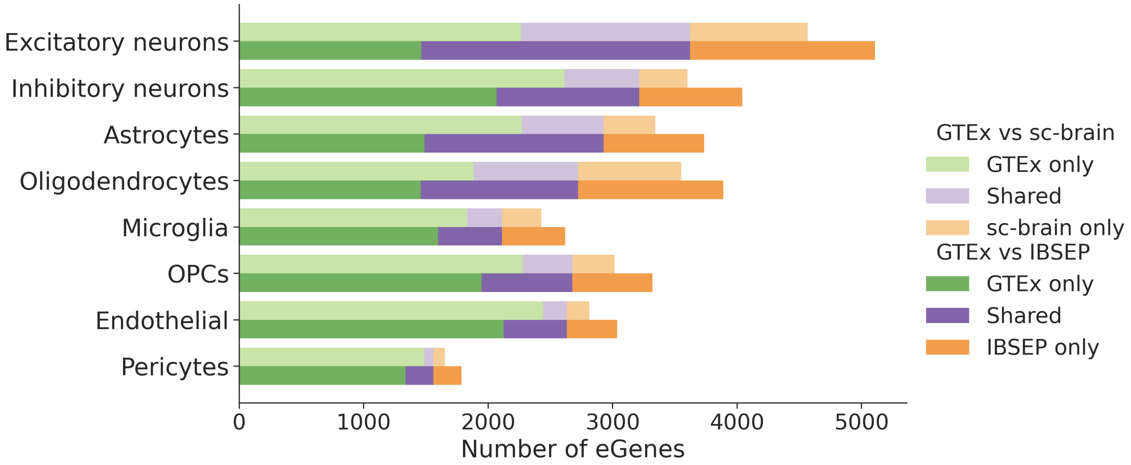

Figure S19: **Barplot comparing the number of eGenes discovered by sc-brain and IBSEP respectively with GTEx in each brain cell type.**

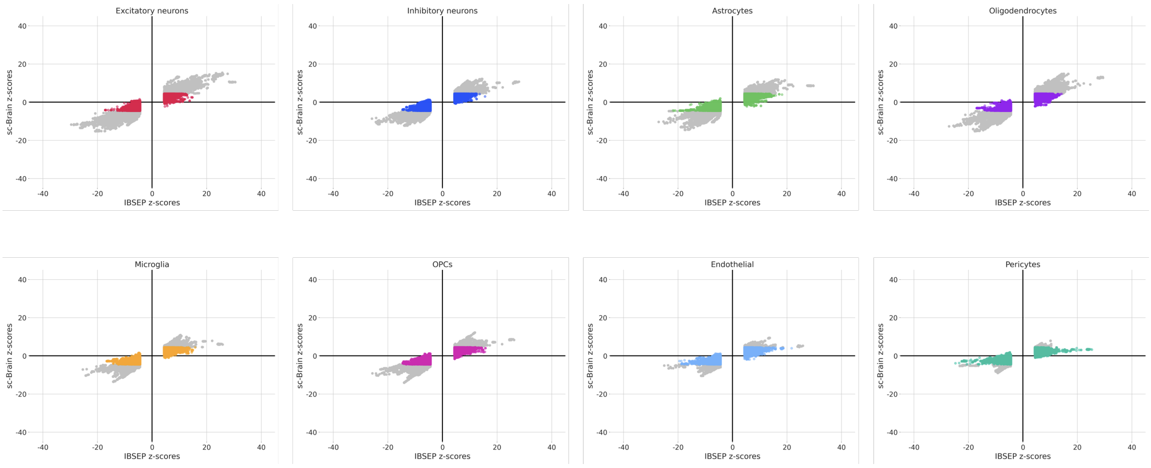

Figure S20: **Comparison of $z$-scores of IBSEP eSNPs with sc-brain.**

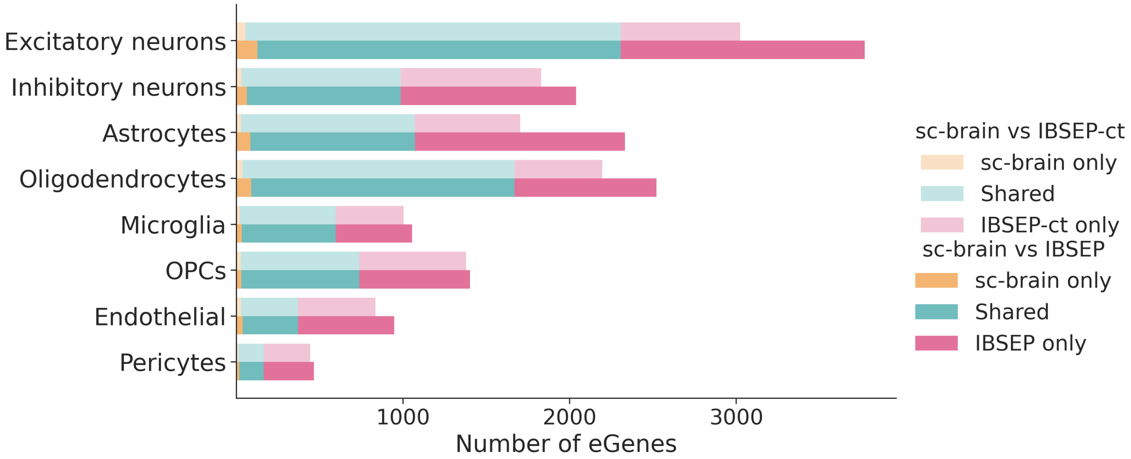

Figure S21: **Barplot comparing the number of eGenes discovered by IBSEP-ct and IBSEP respectively with sc-brain in each brain cell type.**

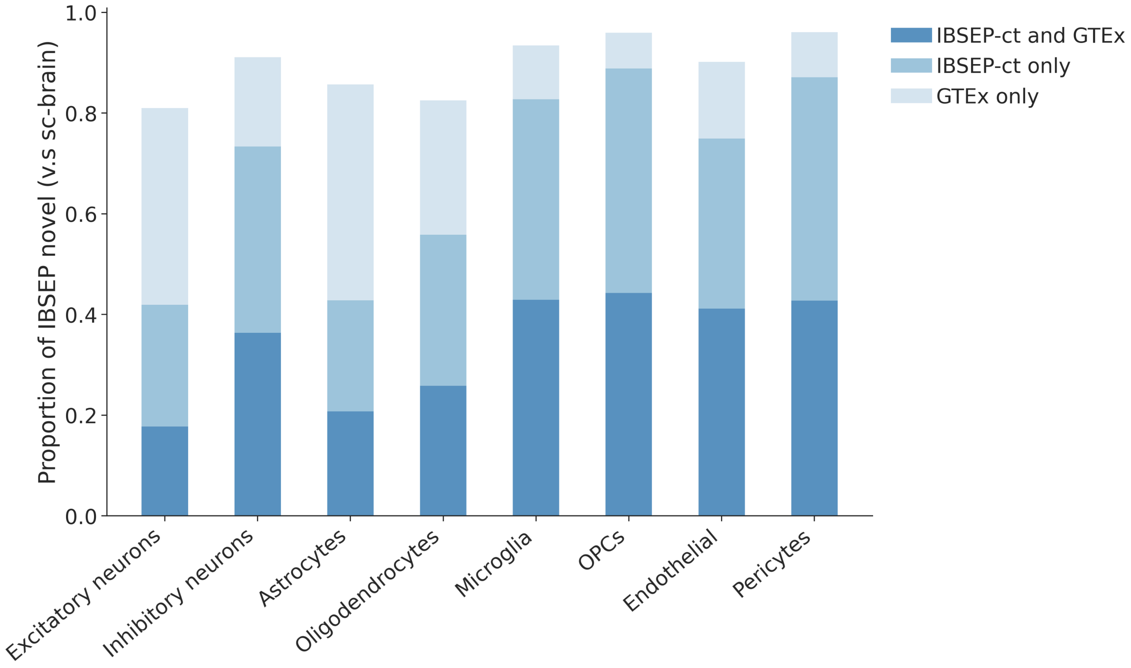

Figure S22: **Proportion of IBSEP novel eGenes shared with GTEx or IBSEP-ct.**

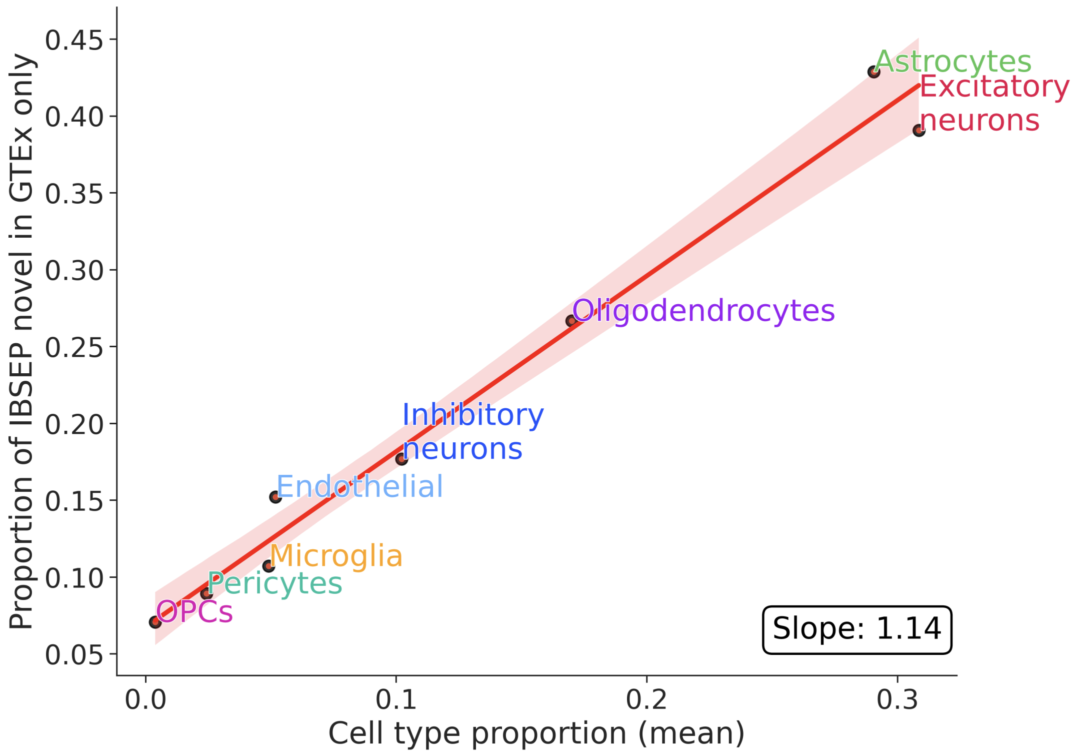

Figure S23: **Relationship between proportion of IBSEP novel eGenes in GTEx only and brain average cell type proportion.**

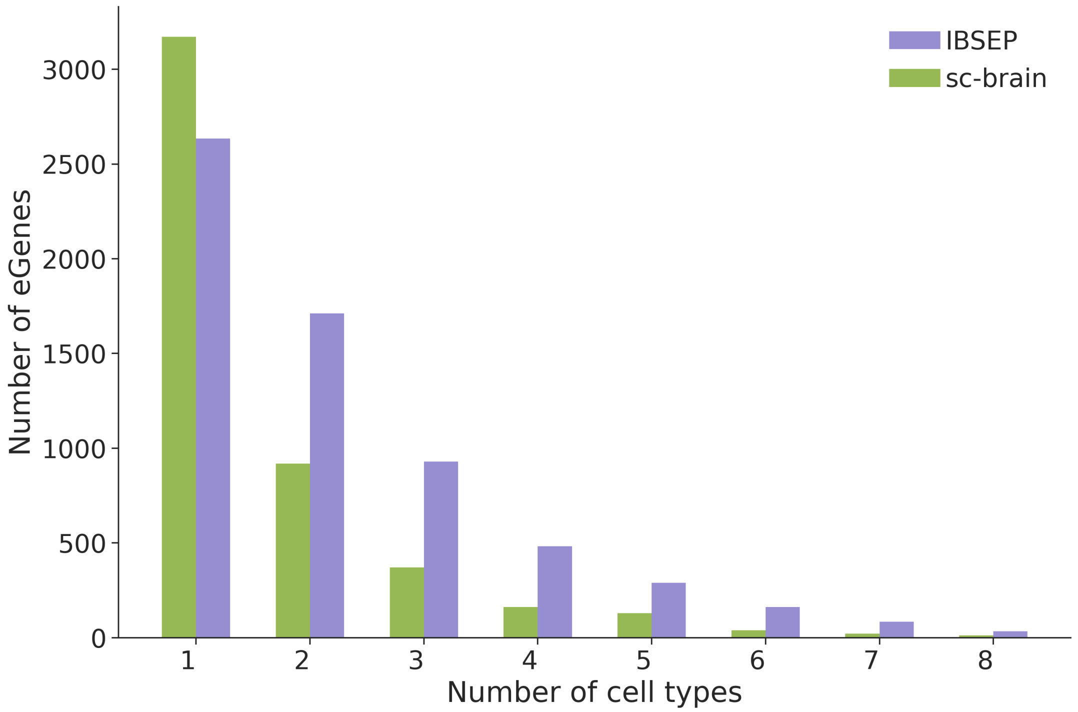

Figure S24: **Number of cell types in which the eGene is identified by sc-brain and IBSEP.**

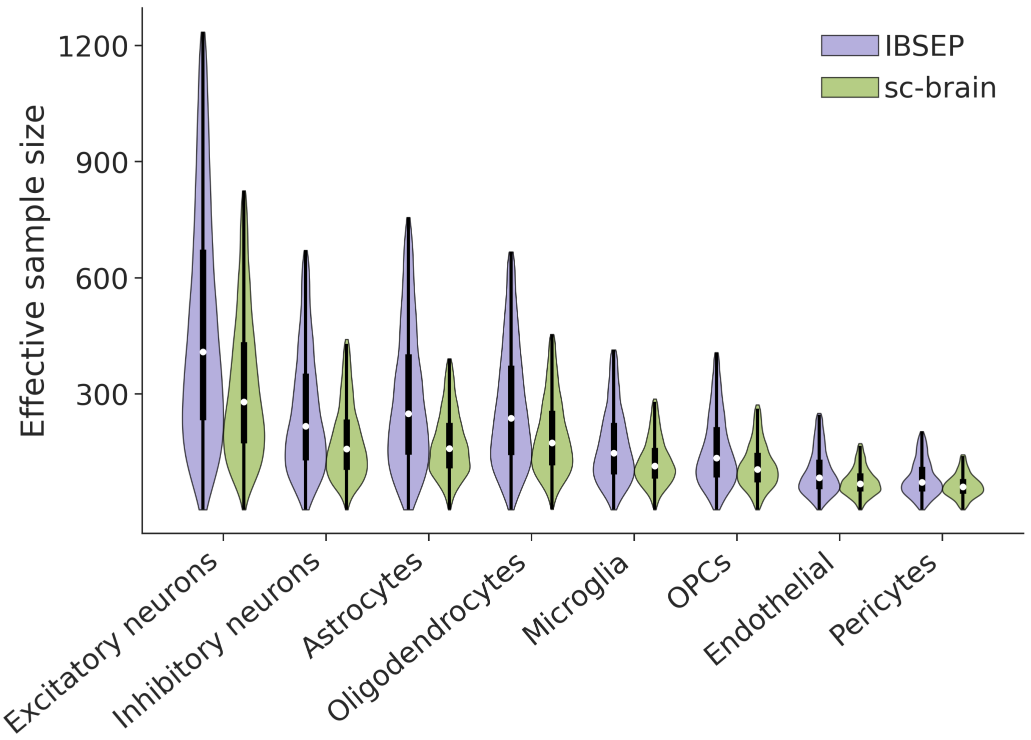

Figure S25: **Distribution of effective sample size of sc-brain and IBSEP for each gene in each cell type.**

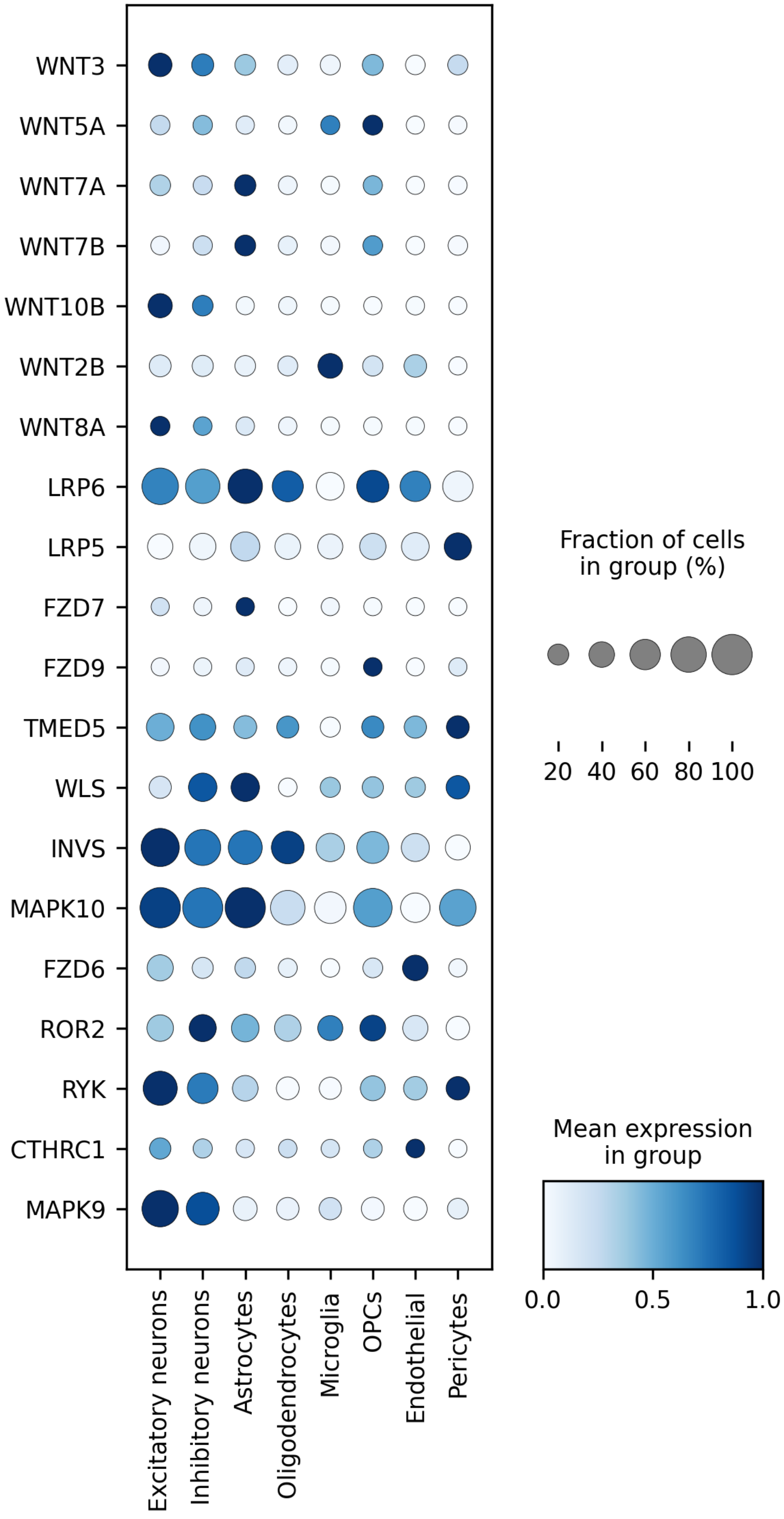

Figure S26: **Expression of Wnt pathway eGenes in a human brain atlas [1].**

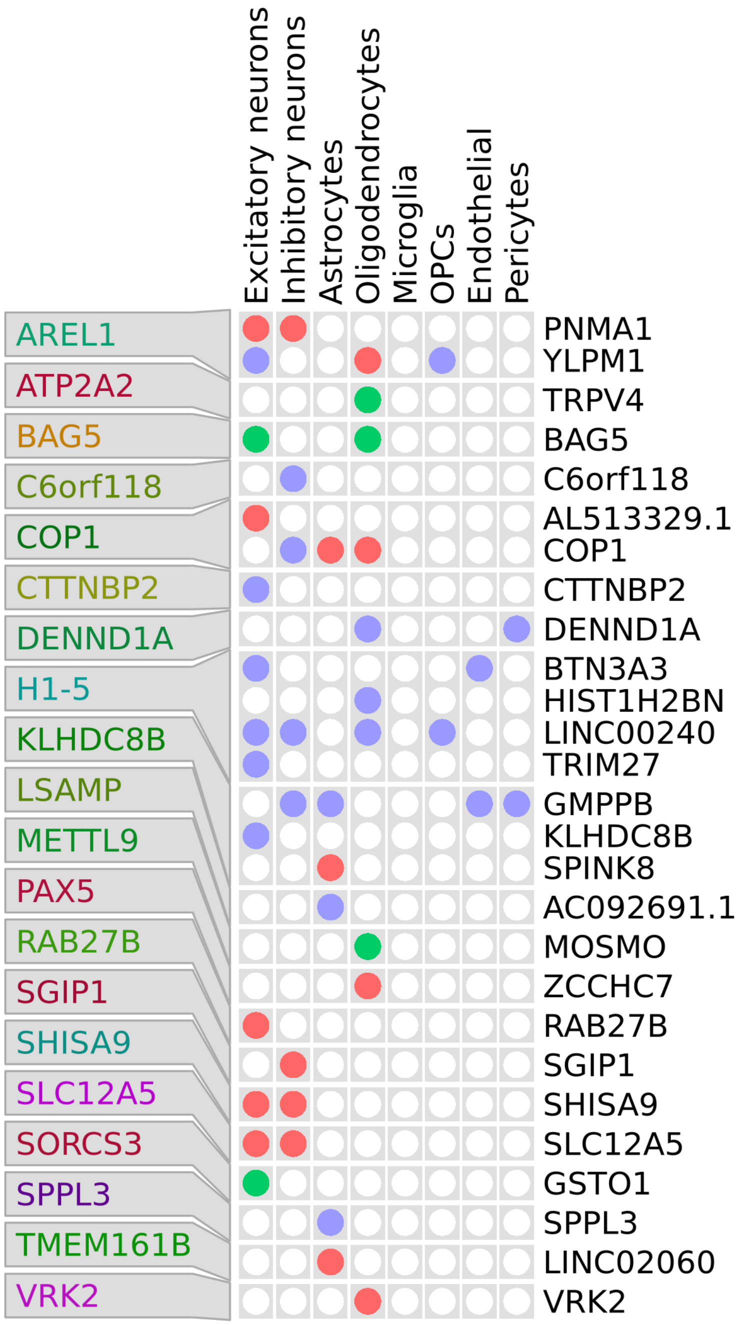

Figure S27: **Cell-type-level colocalized genes of major depressive disease found by sc-brain and IBSEP.**

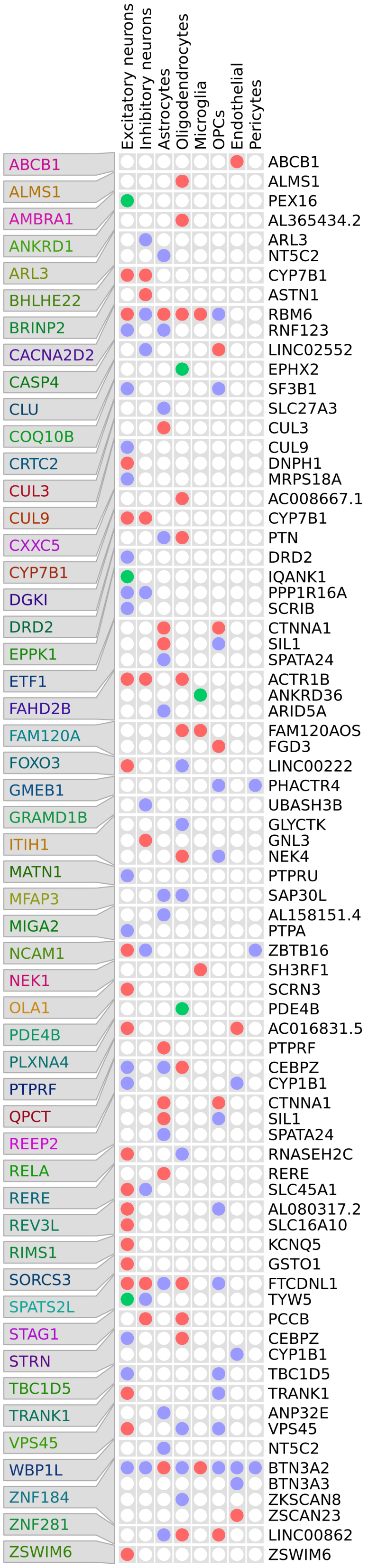

(a) Chromosome 1-11.

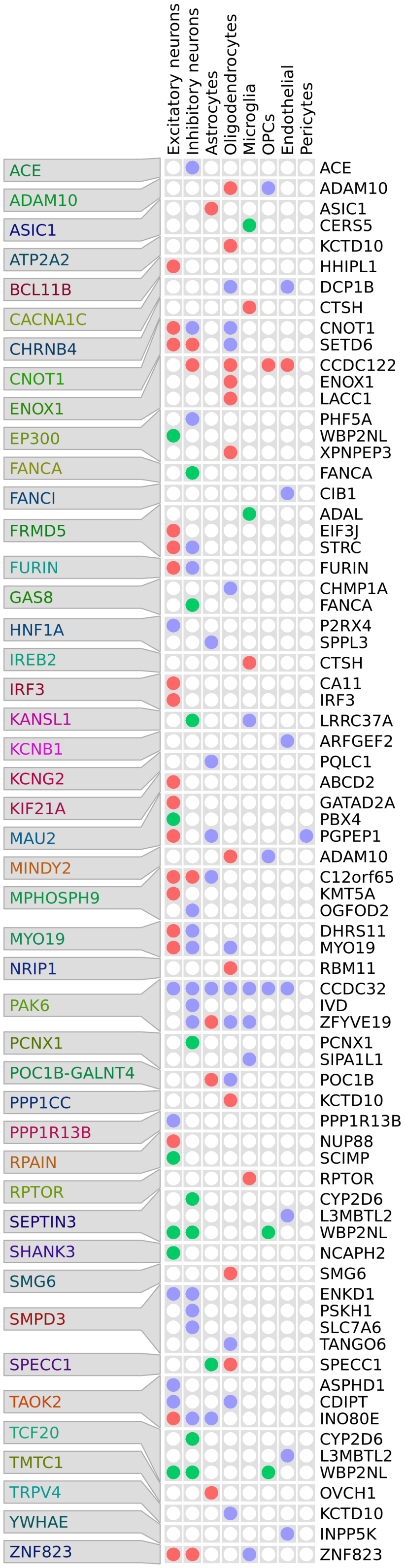

(b) Chromosome 12-22.

Figure S28: **Cell-type-level colocalized genes of schizophrenia found by sc-brain and IBSEP.**

### 2 Supplementary methods

#### 2.1 Derivation of the covariance of marginal effect sizes

We first apply the law of total expectation to obtain the first moments of $\hat{b}_{1j}$, $\hat{b}_{2j}$ and $\hat{b}_{tj}$:

$$\mathbb{E}\left[\hat{b}_{1j}\right] = \mathbb{E}\left[\mathbb{E}\left[\hat{b}_{1j}|\boldsymbol{\beta}_1\right]\right] = 0, \quad \mathbb{E}\left[\hat{b}_{2j}\right] = \mathbb{E}\left[\mathbb{E}\left[\hat{b}_{2j}|\boldsymbol{\beta}_2\right]\right] = 0, \mathbb{E}\left[\hat{b}_{tj}\right] = \mathbb{E}\left[\mathbb{E}\left[\hat{b}_{tj}|\boldsymbol{\beta}_1, \boldsymbol{\beta}_2\right]\right] = 0 \tag{S1}$$

By the law of total variance, we have

$$\begin{aligned}
\mathbb{E}\left[\hat{b}_{1j}^2\right] &= \mathrm{Var}\left[\hat{b}_{1j}\right] \\
&= \mathbb{E}\left[\mathrm{Var}\left[\hat{b}_{1j}|\mathbf{X}_c\right]\right] + \mathrm{Var}\left[\mathbb{E}[\hat{b}_{1j}|\mathbf{X}_c]\right] \\
&= \mathbb{E}\left[\frac{1}{N_c^2}\mathrm{Var}(\mathbf{x}_{cj}^T\mathbf{y}_1|\mathbf{X}_c)\right] + 0 \\
&= \mathbb{E}\left[\sum_{k=1}^{M}(\mathbf{x}_{cj}^T\mathbf{x}_{ck}/N_c)^2\mathrm{Var}(\beta_{1k}) + \frac{1}{N_c^2}\mathrm{Var}(\mathbf{x}_{cj}^T\boldsymbol{\epsilon}_1\mathbf{x}_{cj}|\mathbf{X}_c)\right] \\
&= \omega_1\mathbb{E}\left[\sum_{k=1}^{M}(\mathbf{x}_{cj}^T\mathbf{x}_{ck}/N_c)^2\right] + \frac{1}{N_c}\sigma_{\boldsymbol{\epsilon}_1}^2 \\
&\overset{①}{\approx} \omega_1\sum_{k=1}^{M} r_{jk}^2 + \frac{M\cdot\omega_1}{N_c} + \frac{\sigma_{\boldsymbol{\epsilon}_1}^2}{N_c} \\
&\overset{②}{=} \omega_1\sum_{k=1}^{M} r_{jk}^2 + \frac{1}{N_c} \\
&\overset{③}{\approx} \omega_1 l_j + \hat{s}_{1j}^2,
\end{aligned} \tag{S2}$$

where we use $\mathbb{E}((\mathbf{x}_{cj}^T\mathbf{x}_{ck}/N_c)^2) \approx r_{jk}^2 + 1/N_c$ in approximation ①, $M\cdot\omega_1 + \sigma_{\boldsymbol{\epsilon}_1}^2 = \mathrm{Var}(y_{i1}) = 1$ for equation ②, and approximation ③ is derived from

$$\hat{s}_{1j} = \sqrt{(\mathbf{y}_1 - \mathbf{x}_{cj}\hat{b}_{1j})^T(\mathbf{y}_1 - \mathbf{x}_{cj}\hat{b}_{1j})/(N_c\mathbf{x}_{cj}^T\mathbf{x}_{cj})} \approx \sqrt{\mathbf{y}_1^T\mathbf{y}_1/(N_c\mathbf{x}_{cj}^T\mathbf{x}_{cj})} = 1/\sqrt{N_c}. \tag{S3}$$

Similarly, we can derive $\mathbb{E}\left[\hat{b}_{2j}^2\right] \approx \omega_2 l_j + \hat{s}_{2j}^2$, and

$$\begin{aligned}
\mathbb{E}\left[\hat{b}_{tj}^2\right] &= \mathrm{Var}\left[\hat{b}_{tj}\right] \\
&= \mathbb{E}\left[\mathrm{Var}\left[\hat{b}_{tj}|\mathbf{X}_t\right]\right] + \mathrm{Var}\left[\mathbb{E}[\hat{b}_{tj}|\mathbf{X}_t]\right] \\
&= \mathbb{E}\left[\frac{1}{N_t^2}\mathrm{Var}(\mathbf{x}_{tj}^T\mathbf{y}_t|\mathbf{X}_t)\right] + 0 \\
&\approx \mathbb{E}\left[\sum_{k=1}^{M}(\mathbf{x}_{tj}^T\mathbf{x}_{tk}/N_t)^2\mathrm{Var}(\bar{\pi}_1\beta_{1k} + \bar{\pi}_2\beta_{2k}) + \frac{1}{N_t^2}\mathrm{Var}(\mathbf{x}_{tj}^T\boldsymbol{\epsilon}_t\mathbf{x}_{tj}|\mathbf{X}_t)\right] \\
&= (\bar{\pi}_1^2\omega_1 + \bar{\pi}_2^2\omega_2 + \bar{\pi}_1\bar{\pi}_2\omega_{12})\mathbb{E}\left[\sum_{k=1}^{M}(\mathbf{x}_{tj}^T\mathbf{x}_{tk}/N_t)^2\right] + \frac{1}{N_t}\sigma_{\boldsymbol{\epsilon}_t}^2 \\
&\approx (\bar{\pi}_1^2\omega_1 + \bar{\pi}_2^2\omega_2 + \bar{\pi}_1\bar{\pi}_2\omega_{12})\sum_{k=1}^{M}r_{jk}^2 + \frac{M}{N_t}(\bar{\pi}_1^2\omega_1 + \bar{\pi}_2^2\omega_2 + \bar{\pi}_1\bar{\pi}_2\omega_{12}) + \frac{\sigma_{\boldsymbol{\epsilon}_t}^2}{N_t} \\
&= (\bar{\pi}_1^2\omega_1 + \bar{\pi}_2^2\omega_2 + \bar{\pi}_1\bar{\pi}_2\omega_{12})\sum_{k=1}^{M}r_{jk}^2 + \frac{1}{N_t} \\
&\approx (\bar{\pi}_1^2\omega_1 + \bar{\pi}_2^2\omega_2 + \bar{\pi}_1\bar{\pi}_2\omega_{12})l_j + \hat{s}_{tj}^2,
\end{aligned} \tag{S4}$$

The covariance between $\hat{b}_{1j}$ and $\hat{b}_{2j}$ can be obtained as

$$\begin{aligned}
\mathrm{Cov}\left[\hat{b}_{1j}, \hat{b}_{2j}\right] &= \mathbb{E}\left[\hat{b}_{1j}\hat{b}_{2j}\right] \\
&= \frac{1}{N_c^2}\mathbb{E}\left[\mathbb{E}\left[\mathbf{x}_{cj}^T\mathbf{y}_1\mathbf{y}_2^T\mathbf{x}_{cj}|\mathbf{X}_c\right]\right] \\
&= \frac{1}{N_c^2}\mathbb{E}\left[\mathbf{x}_{cj}^T\mathbb{E}\left[(\mathbf{X}_c\boldsymbol{\beta}_1 + \boldsymbol{\epsilon}_1)(\mathbf{X}_c\boldsymbol{\beta}_2 + \boldsymbol{\epsilon}_2)^T|\mathbf{X}_c\right]\mathbf{x}_{cj}\right] \\
&= \frac{1}{N_c^2}\mathbb{E}\left[\mathbf{x}_{cj}^T\mathbf{X}_c\mathbb{E}(\boldsymbol{\beta}_1\boldsymbol{\beta}_2^T)\mathbf{X}_c^T\mathbf{x}_{cj}\right] \\
&= \omega_{12}\mathbb{E}\left[\sum_{k=1}^{M}(\mathbf{x}_{cj}^T\mathbf{x}_{ck}/N_c)^2\right] \\
&= \omega_{12}\sum_{k=1}^{M}r_{jk}^2 \\
&= \omega_{12}l_j,
\end{aligned} \tag{S5}$$

Similarly,

$$
\begin{aligned}
\text{Cov}\left[\hat{b}_{1j},\hat{b}_{tj}\right] &= \mathbb{E}\left[\hat{b}_{1j}\hat{b}_{tj}\right] \\
&= \frac{1}{N_cN_t}\mathbb{E}\left[\mathbb{E}\left[\mathbf{x}_{cj}^T\mathbf{y}_1\mathbf{y}_t^T\mathbf{x}_{tj}|\mathbf{X}_c,\mathbf{X}_t\right]\right] \\
&\approx \frac{1}{N_cN_t}\mathbb{E}\left[\mathbf{x}_{cj}^T\mathbb{E}\left[(\mathbf{X}_c\boldsymbol{\beta}_1+\boldsymbol{\epsilon}_1)(\mathbf{X}_t(\bar{\pi}_1\boldsymbol{\beta}_1+\bar{\pi}_2\boldsymbol{\beta}_2)+\boldsymbol{\epsilon}_t)^T|\mathbf{X}_c,\mathbf{X}_t\right]\mathbf{x}_{tj}\right] \\
&= \frac{1}{N_cN_t}\mathbb{E}\left[\mathbf{x}_{cj}^T\mathbf{X}_c\mathbb{E}(\boldsymbol{\beta}_1(\bar{\pi}_1\boldsymbol{\beta}_1+\bar{\pi}_2\boldsymbol{\beta}_2)^T)\mathbf{X}_t^T\mathbf{x}_{tj}\right] \\
&= (\bar{\pi}_1\omega_1+\bar{\pi}_2\omega_{12})\mathbb{E}\left[\sum_{k=1}^{M}(\mathbf{x}_{cj}^T\mathbf{x}_{tk})^2/(N_cN_t)\right] \\
&= (\bar{\pi}_1\omega_1+\bar{\pi}_2\omega_{12})\sum_{k=1}^{M}r_{jk}^2 \\
&= (\bar{\pi}_1\omega_1+\bar{\pi}_2\omega_{12})l_j,
\end{aligned}
\tag{S6}
$$

and similarly $\text{Cov}\left[\hat{b}_{2j},\hat{b}_{tj}\right]=(\bar{\pi}_1\omega_{12}+\bar{\pi}_2\omega_2)l_j$.

#### 2.2 Covariance of marginal effect sizes accounting for population stratification

We model population stratification following LDSC [2] and derive the covariance of marginal effect sizes in a same way as our previous work [3]. The final expressions of covariances are similar to those free of population stratification but with extra coefficients and constants:

$$
\begin{aligned}
\mathbb{E}[\hat{b}_{1j}] &= \mathbb{E}[\hat{b}_{2j}] = \mathbb{E}[\hat{b}_{tj}] = 0, \\
\mathbb{E}[\hat{b}_{1j}^2] &= \omega_1 l_j + c_1\hat{s}_{1j}^2, \\
\mathbb{E}[\hat{b}_{2j}^2] &= \omega_2 l_j + c_2\hat{s}_{2j}^2, \\
\mathbb{E}[\hat{b}_{tj}^2] &= (\bar{\pi}_1^2\omega_1+\bar{\pi}_2^2\omega_2+2\bar{\pi}_1\bar{\pi}_2\omega_{12})l_j + c_3\hat{s}_{tj}^2, \\
\text{Cov}[\hat{b}_{1j},\hat{b}_{2j}] &= \omega_{12}l_j + c_{12}\hat{s}_{1j}\hat{s}_{2j}, \\
\text{Cov}[\hat{b}_{1j},\hat{b}_{tj}] &= (\bar{\pi}_1\omega_1+\bar{\pi}_2\omega_{12})l_j + c_{13}\hat{s}_{1j}\hat{s}_{tj}, \\
\text{Cov}[\hat{b}_{2j},\hat{b}_{tj}] &= (\bar{\pi}_1\omega_{12}+\bar{\pi}_2\omega_2)l_j + c_{23}\hat{s}_{2j}\hat{s}_{tj}.
\end{aligned}
\tag{S7}
$$

where $c_1, c_2, c_3, c_{12}, c_{13}, c_{23}$ are inflation constants that adjust for the confounding biases due to the geographic structure or sample overlap etc.

#### 2.3 Theoretical justification on the usage of mean cell type proportions

Here we show that under some mild conditions, the marginal effect size $\hat{b}_{tj}$ for cis-SNP $j$ in tissue eQTL data can be approximated by that computed from the tissue-level eQTL model with average cell type proportions:

$$
\begin{aligned}
\hat{b}_{tj} &= \mathbf{x}_{tj}^T \mathbf{y}_t / \mathbf{x}_{tj}^T \mathbf{x}_{tj} \\
&= \mathbf{x}_{tj}^T \mathbf{y}_t / N_t \\
&= \mathbf{x}_{tj}^T (\boldsymbol{\pi}_1 \mathbf{X}_t \boldsymbol{\beta}_1 + \boldsymbol{\pi}_2 \mathbf{X}_t \boldsymbol{\beta}_2 + \boldsymbol{\epsilon}_t) / N_t \\
&= \sum_{i=1}^{N_t} x_{tj,i} (\pi_{1i} \mathbf{x}_{t,i} \boldsymbol{\beta}_1 + \pi_{2i} \mathbf{x}_{t,i} \boldsymbol{\beta}_2 + \boldsymbol{\epsilon}_{t,i}) / N_t \\
&= \sum_{i=1}^{N_t} x_{tj,i} \left[ \pi_{1i} (x_{tj,i} \beta_{1j} + \sum_{j'=1, j' \neq j}^{M} x_{tj',i} \beta_{1j'}) + \pi_{2i} (x_{tj,i} \beta_{1j} + \sum_{j'=1, j' \neq j}^{M} x_{tj',i} \beta_{1j'}) + \boldsymbol{\epsilon}_{t,i} \right] / N_t \\
&\overset{①}{=} \sum_{i=1}^{N_t} x_{tj,i} (\pi_{1i} x_{tj,i} \beta_{1j} + \pi_{2i} x_{tj,i} \beta_{1j}) / N_t + const. \\
&= \sum_{i=1}^{N_t} (\pi_{1i} x_{tj,i}^2 \beta_{1j} + \pi_{2i} x_{tj,i}^2 \beta_{1j}) / N_t + const. \\
&= ( \sum_{g_{tj,i}=0} (\pi_{1i} x_{tj,i}^2 \beta_{1j} + \pi_{2i} x_{tj,i}^2 \beta_{1j}) + \sum_{g_{tj,i}=1} (\pi_{1i} x_{tj,i}^2 \beta_{1j} + \pi_{2i} x_{tj,i}^2 \beta_{1j}) + \sum_{g_{tj,i}=2} (\pi_{1i} x_{tj,i}^2 \beta_{1j} + \pi_{2i} x_{tj,i}^2 \beta_{1j})) / N_t + const. \\
&\overset{②}{\approx} ( \sum_{g_{tj,i}=0} (\bar{\pi}_1 x_{tj,i}^2 \beta_{1j} + \bar{\pi}_2 x_{tj,i}^2 \beta_{1j}) + \sum_{g_{tj,i}=1} (\bar{\pi}_1 x_{tj,i}^2 \beta_{1j} + \bar{\pi}_2 x_{tj,i}^2 \beta_{1j}) + \sum_{g_{tj,i}=2} (\bar{\pi}_1 x_{tj,i}^2 \beta_{1j} + \bar{\pi}_2 x_{tj,i}^2 \beta_{1j})) / N_t + const. \\
&= \sum_{i=1}^{N_t} (\bar{\pi}_1 x_{tj,i}^2 \beta_{1j} + \bar{\pi}_2 x_{tj,i}^2 \beta_{1j}) / N_t + const. \\
&\overset{③}{\approx} \sum_{i=1}^{N_t} x_{tj,i} (\bar{\pi}_1 \mathbf{x}_{t,i} \boldsymbol{\beta}_1 + \bar{\pi}_2 \mathbf{x}_{t,i} \boldsymbol{\beta}_2 + \boldsymbol{\epsilon}_{t,i}) / N_t \\
&= \mathbf{x}_{tj}^T (\bar{\pi}_1 \mathbf{X}_t \boldsymbol{\beta}_1 + \bar{\pi}_2 \mathbf{X}_t \boldsymbol{\beta}_2 + \boldsymbol{\epsilon}_t) / N_t
\end{aligned} \tag{S8}
$$

where $g_{tj,i}$ is the raw genotype corresponding to $x_{tj,i}$, $\bar{\pi}_1$ and $\bar{\pi}_2$ are mean cell type proportions across individuals in the tissue eQTL data. We consider SNP $j$ only in ①, and we use the law of large number in approximation ② when $N_t$ is large enough. Similarly, we can derive approximation ③ using the same idea. In conclusion, the marginal effect size $\hat{b}_{tj}$ can be approximately estimated using mean cell type proportions.

#### 2.4 Derivation of IBSEP estimator

IBSEP estimator is derived in the framework of GMM which requires moment conditions and a weight matrix. The moment conditions are the following linear projections:

$$\mathbb{E}\left[\hat{b}_{2j} - b_{1j}\xi\right] = 0, \tag{S9}$$

$$\mathbb{E}\left[\hat{b}_{tj} - b_{1j}\gamma\right] = 0, \tag{S10}$$

where $\xi, \gamma$ are unknown parameters. In GMM, $\xi, \gamma$ are derived by minimizing the objective functions, respectively:

$$\mathbb{E}\left[(\hat{b}_{2j} - b_{1j}\xi)^2\right], \tag{S11}$$

$$\mathbb{E}\left[(\hat{b}_{tj} - b_{1j}\gamma)^2\right]. \tag{S12}$$

Set the derivative of Eq. S11 w.r.t $\xi$ and derivatives of Eq. S12 w.r.t $\gamma_1, \gamma_2$ to be zero:

$$\begin{aligned}
0 &= \frac{\partial}{\partial \xi}\mathbb{E}\left[(\hat{b}_{2j} - b_{1j}\xi)^2\right] \\
&= \mathbb{E}\left[\frac{\partial}{\partial \xi}(\hat{b}_{2j} - b_{1j}\xi)^2\right] \\
&= 2\mathbb{E}\left[b_{1j}(\hat{b}_{2j} - b_{1j}\xi)\right] \\
&= 2\mathbb{E}\left[b_{1j}(b_{2j} + e_{2j} - b_{1j}\xi)\right] \\
&= 2\mathbb{E}\left[b_{1j}b_{2j}\right] - 2\mathbb{E}\left[b_{1j}b_{1j}\xi\right] \\
&= 2\omega_{j,12} - 2\omega_{j,11}\xi,
\end{aligned} \tag{S13}$$

$$\begin{aligned}
0 &= \frac{\partial}{\partial \gamma}\mathbb{E}\left[(\hat{b}_{tj} - b_{1j}\gamma)^2\right] \\
&= \mathbb{E}\left[\frac{\partial}{\partial \gamma}(\hat{b}_{tj} - b_{1j}\gamma)^2\right] \\
&= 2\mathbb{E}\left[b_{1j}(\hat{b}_{tj} - b_{1j}\gamma)\right] \\
&= 2\mathbb{E}\left[b_{1j}(\bar{\pi}_1 b_{1j} + \bar{\pi}_2 b_{2j} + e_{tj} - b_{1j}\gamma)\right] \\
&= 2\bar{\pi}_1\mathbb{E}\left[b_{1j}b_{2j}\right] + 2\bar{\pi}_2\mathbb{E}\left[b_{1j}b_{2j}\right] - 2\mathbb{E}\left[b_{1j}b_{1j}\gamma\right] \\
&= 2\bar{\pi}_1\omega_{j,11} + 2\bar{\pi}_2\omega_{j,12} - 2\omega_{j,11}\gamma,
\end{aligned} \tag{S14}$$

then

$$\xi = \frac{\omega_{j,12}}{\omega_{j,11}}, \tag{S15}$$

$$\gamma = \bar{\pi}_1 + \bar{\pi}_2\frac{\omega_{j,12}}{\omega_{j,11}}. \tag{S16}$$

We have shown that the projections of $\hat{b}_{2j}$ and $\hat{b}_{tj}$ onto $b_{1j}$ are $\frac{\omega_{j,12}}{\omega_{j,11}}b_{1j}$ and $\left(\bar{\pi}_1 + \bar{\pi}_2\frac{\omega_{j,12}}{\omega_{j,11}}\right)b_{1j}$, respectively. Hence we obtain the conditional mean of the estimated marginal effects:

$$\mathbb{E}\left[\begin{pmatrix}\hat{b}_{1j}\\ \hat{b}_{2j}\\ \hat{b}_{tj}\end{pmatrix}|b_{1j}\right] = \underbrace{\begin{pmatrix}1\\ \omega_{j,12}/\omega_{j,11}\\ \bar{\pi}_1 + \bar{\pi}_2\omega_{j,12}/\omega_{j,11}\end{pmatrix}}_{\boldsymbol{\lambda}_1} b_{1j}, \tag{S17}$$

Moreover, we can derive the conditional variance:

$$\begin{aligned}
\mathrm{Var}\left[\begin{pmatrix}\hat{b}_{1j}\\ \hat{b}_{2j}\\ \hat{b}_{tj}\end{pmatrix}|b_{1j}\right] &= \mathrm{Var}\left[\begin{pmatrix}b_{1j}\\ b_{2j}\\ b_{tj}\end{pmatrix}|b_{1j}\right] + \mathrm{Var}\left[\begin{pmatrix}e_{1j}\\ e_{2j}\\ e_{tj}\end{pmatrix}|b_{1j}\right] \\
&= \mathbf{A}\left(\boldsymbol{\Omega}_j - \frac{\boldsymbol{\omega}_{j,\cdot 1}\boldsymbol{\omega}_{j,\cdot 1}^T}{\omega_{j,11}}\right)\mathbf{A}^T + \hat{\mathbf{S}}_j\mathbf{C}\hat{\mathbf{S}}_j := \boldsymbol{\Lambda}_1^{-1}.
\end{aligned} \tag{S18}$$

Now, using $\boldsymbol{\Lambda}_1$ as the weight matrix, we can obtain a GMM estimator of $b_{1j}$ by minimizing

$$\mathbf{m}(b)^T\boldsymbol{\Lambda}_1\mathbf{m}(b) = \left[\begin{pmatrix}\hat{b}_{1j}\\ \hat{b}_{2j}\\ \hat{b}_{tj}\end{pmatrix} - \boldsymbol{\lambda}_1 b_{1j}\right]^T \boldsymbol{\Lambda}_1 \left[\begin{pmatrix}\hat{b}_{1j}\\ \hat{b}_{2j}\\ \hat{b}_{tj}\end{pmatrix} - \boldsymbol{\lambda}_1 b_{1j}\right]. \tag{S19}$$

By setting the derivative of this objective w.r.t $b_{1j}$ to be zero:

$$\begin{aligned} 0 &= \frac{\partial}{\partial b_{1j}} \left[\mathbf{m}(b)^T \mathbf{\Lambda}_1 \mathbf{m}(b)\right] \\ &= 2\boldsymbol{\lambda}_1^T \mathbf{\Lambda} \boldsymbol{\lambda} b_{1j} - 2\boldsymbol{\lambda}^T \mathbf{\Lambda} \begin{pmatrix} \hat{b}_{1j} \\ \hat{b}_{2j} \\ \hat{b}_{tj} \end{pmatrix}, \end{aligned} \tag{S20}$$

we have

$$\hat{b}_{1j}^{\text{IBSEP}} = \arg\min_b \mathbf{m}(b)^T \mathbf{\Lambda}_1 \mathbf{m}(b) = \underbrace{\left(\boldsymbol{\lambda}_1^T \mathbf{\Lambda}_1 \boldsymbol{\lambda}_1\right)^{-1} \boldsymbol{\lambda}_1^T \mathbf{\Lambda}_1}_{\mathbf{w}_1^T} \underbrace{\begin{pmatrix} \hat{b}_{1j} \\ \hat{b}_{2j} \\ \hat{b}_{tj} \end{pmatrix}}_{\hat{\mathbf{b}}_{\cdot,j}} = \mathbf{w}_1^T \hat{\mathbf{b}}_{\cdot,j}. \tag{S21}$$

#### 2.5 Theoretical properties of IBSEP estimator

Following previous study [3], we can show that IBSEP estimator is the best linear unbiased estimator (BLUE). To see this, we first show the IBSEP estimate (e.g., $\hat{b}_{1j}^{\text{IBSEP}}$) is an unbiased estimator of the true marginal effect $b_{1j}$:

$$\mathbb{E}\left[\hat{b}_{1j}^{\text{IBSEP}} | b_{1j}\right] = \mathbb{E}\left[\mathbf{w}_1^T \hat{\mathbf{b}}_{\cdot,j} | b_{1j}\right] = \mathbf{w}_1^T \mathbb{E}\left[\hat{\mathbf{b}}_{\cdot,j} | b_{1j}\right] \overset{\text{Eq.}(S17)}{=} \mathbf{w}_1^T \frac{\boldsymbol{\omega}_{j,\cdot 1}}{\omega_{j,11}} b_{1j} = b_{1j}. \tag{S22}$$

Next, we consider a necessary condition for the set of all unbiased linear estimators denoted as $(\mathbf{w}_1 + \mathbf{u})^T \hat{\mathbf{b}}_{\cdot,j}$, where $\mathbf{u}$ represents some vectors such that $\mathbb{E}\left[(\mathbf{w}_1 + \mathbf{u})^T \hat{\mathbf{b}}_{\cdot,j} | b_{1j}\right] = b_{1j}$. It follows that

$$\begin{aligned} 0 &= \mathbb{E}\left[(\mathbf{w}_1 + \mathbf{u})^T \hat{\mathbf{b}}_{\cdot,j} | b_{1j}\right] - b_{1j} \\ &= \mathbb{E}\left[\mathbf{w}_1^T \hat{\mathbf{b}}_{\cdot,j} | b_{1j}\right] + \mathbb{E}\left[\mathbf{u}^T \hat{\mathbf{b}}_{\cdot,j} | b_{1j}\right] - b_{1j} \\ &= b_{1j} + \mathbb{E}\left[\mathbf{u}^T \hat{\mathbf{b}}_{\cdot,j} | b_{1j}\right] - b_{1j} \text{ (by the unbiasedness of IBSEP)} \\ &= \mathbb{E}\left[\mathbf{u}^T \hat{\mathbf{b}}_{\cdot,j} | b_{1j}\right] \\ &= \mathbf{u}^T \frac{\boldsymbol{\omega}_{j,\cdot 1}}{\omega_{j,11}} b_{1j}. \end{aligned} \tag{S23}$$

Therefore, we obtain $\mathbf{u}^T \boldsymbol{\omega}_{j,\cdot 1} = 0$. We then show that the variance $\text{Var}\left[(\mathbf{w}_1 + \mathbf{u})^T \hat{\mathbf{b}}_{\cdot,j} | b_{1j}\right]$ is larger than the variance $\text{Var}\left[\mathbf{w}_1^T \hat{\mathbf{b}}_{\cdot,j} | b_{1j}\right]$. We expand

$$\text{Var}\left[(\mathbf{w}_1 + \mathbf{u})^T \hat{\mathbf{b}}_{\cdot,j} | b_{1j}\right] = \mathbf{w}_1^T \text{Var}(\hat{\mathbf{b}}_{\cdot,j} | b_{1j}) \mathbf{w}_1 + \mathbf{u}^T \text{Var}(\hat{\mathbf{b}}_{\cdot,j} | b_{1j}) \mathbf{u} + 2\mathbf{w}_1^T \text{Var}(\hat{\mathbf{b}}_{\cdot,j} | b_{1j}) \mathbf{u}. \tag{S24}$$

Based on the asymptotic normality of GMM, we have

$$\begin{aligned} \mathbf{w}_1^T \text{Var}(\hat{\mathbf{b}}_{\cdot,j} | b_{1j}) \mathbf{u} &= \left(\frac{\boldsymbol{\omega}_{j,\cdot 1}^T}{\omega_{j,11}} \mathbf{\Lambda}_1 \frac{\boldsymbol{\omega}_{j,\cdot 1}}{\omega_{j,11}}\right)^{-1} \frac{\boldsymbol{\omega}_{j,\cdot 1}^T}{\omega_{j,11}} \mathbf{\Lambda}_1 \mathbf{\Lambda}_1^{-1} \mathbf{u} \\ &= \left(\frac{\boldsymbol{\omega}_{j,\cdot 1}^T}{\omega_{j,11}} \mathbf{\Lambda}_1 \frac{\boldsymbol{\omega}_{j,\cdot 1}}{\omega_{j,11}}\right)^{-1} \frac{\boldsymbol{\omega}_{j,\cdot 1}^T}{\omega_{j,11}} \mathbf{u} \overset{\text{Eq.}(S23)}{=} 0. \end{aligned} \tag{S25}$$

Therefore, we have $\text{Var}\left[(\mathbf{w}_1+\mathbf{u})^T\hat{\mathbf{b}}_{\cdot,j}|b_{1j}\right] = \mathbf{w}_1^T\text{Var}(\hat{\mathbf{b}}_{\cdot,j}|b_{1j})\mathbf{w}_1 + \mathbf{u}^T\text{Var}(\hat{\mathbf{b}}_{\cdot,j}|b_{1j})\mathbf{u} \geq \text{Var}\left[\mathbf{w}_1^T\hat{\mathbf{b}}_{\cdot,j}|b_{1j}\right] = \mathbf{w}_1^T\text{Var}(\hat{\mathbf{b}}_{\cdot,j}|b_{1j})\mathbf{w}_1$. Finnaly, we demonstrate that IBSEP has the lowest variance among all the unbiased estimators.
